# A Deep Ensemble Encoder Network Method for Improved Polygenic Risk Score Prediction

**DOI:** 10.1101/2024.07.31.24311311

**Authors:** Okan Bilge Ozdemir, Raelynn Chen, Ruowang Li

**Affiliations:** Cedars-Sinai Medical Center, Computational Biology Department, Los Angeles, CA, USA

**Keywords:** Genomics, PRS, Deep Learning, Autoencoders

## Abstract

Genome-wide association studies of various heritable human traits and diseases have identified numerous associated single nucleotide polymorphisms (SNPs), most of which have small or modest effects. Polygenic risk scores aim to better estimate individuals’ genetic predisposition by aggregating the effects of multiple SNPs from GWAS. However, current PRS is designed to capture only simple linear genetic effects across the genome, limiting their ability to fully account for the complex polygenic architecture. To address this, we propose DeepEnsembleEncodeNet (DEEN), a new method that ensembles autoencoders and fully connected neural networks to better identify and model linear and non-linear SNP effects across different genomic regions, improving its ability to predict disease risks. To demonstrate DEEN’s performance, we optimized the model across binary and continuous traits from the UK Biobank. Model evaluation on the held-out UK Biobank testing dataset, as well as the independent All of Us dataset, showed improved prediction and risk stratification, consistently outperforming other methods.

## Introduction

Many human traits and diseases are highly heritable, reflecting the important influences of the underlying genetics^1–5^. To date, Genome-wide association studies (GWAS) studies have identified over 70,000 SNP associations spanning a wide range of human traits and diseases^6–11^. Effective leveraging of these genotype-phenotype correlations to construct genetic risk prediction models holds substantial clinical promise by enabling early and stable risk predictions^12–14^. However, individual SNPs typically account for only a fraction of phenotype variability. The recent development of polygenic risk score^15–17^ (PRS), which aggregates univariate effects from many genetic loci identified through GWAS, has shown improved performance in predicting and stratifying genetic susceptibilities in large populations^18–20^. Nevertheless, existing PRS methodologies are constrained by inflexible underlying assumptions of genetic data that limit their ability to fully capture the predictive signals^21^.

Broadly speaking, existing PRS methodologies, such as pruning and thresholding^22–24^, Lasso regularized regressions^25^ or Bayesian methods^26–30^, vary in their approaches to estimate the effects of individual SNPs. Nonetheless, these methodologies often share similar fundamental assumptions. Current PRS models primarily focus on the effects of univariate SNPs and their linear additive aggregations, thus not allowing potential non-linear effects to be captured^31,32^. In addition, they also generally assume uniform signal distributions across the genome, as reflected by fixed modeling parameters, e.g., a single *λ*in Lasso, applied to all SNPs. Furthermore, given the high dimensionality and sparsity of genetic data, many PRS approaches utilize dimensionality reduction or variable selection techniques to improve the signal-to-noise ratio in the input feature space. However, dimensionality reduction typically occurs concurrently with classification or regression in the supervised learning setting. Separating these tasks could yield more efficient methods, as better strategies exist for each task independently.

Autoencoders are highly efficient in data dimensionality reduction, making them particularly valuable in areas such as imaging and natural language processing^33–37^. They function by learning a lower-dimensional representation of the data that can minimize data reconstruction error, effectively capturing key features while discarding noise and redundant information. On the other hand, fully connected neural networks (FCNN) are considered state-of-the-art in predictive modeling due to their ability to process complex, high-dimensional data and learn complex patterns^38–41^. The architecture of FCNN allows for the learning of hierarchical representations of underlying data, improving tasks such as regression and classification. However, despite their respective strengths, autoencoders and FCNNs have not been utilized in ensemble learning to integrate latent representation learning with predictive modeling for improving genetic risk prediction models^42–44^.

In this study, we propose a novel method, DeepEnsembleEncodeNet (DEEN), which utilizes an autoencoder for learning latent genetic feature representations coupled with a FCNN for constructing predictive models. DEEN disentangles genetic data dimensionality reduction and prediction model construction into separate modules, allowing optimal learning for each task. The autoencoder module extracts a lower-dimensional latent representation of the genetic data that can capture both linear and non-linear relationships among the SNPs. Subsequently, the FCNN further enables learning of non-linear effects as well as differential variable weighting across the genome, providing a substantially more flexible framework for capturing diverse genetic effects in constructing genetic risk prediction models. We optimized DEEN using binary diseases, type 2 diabetes (T2D) and hypertension, and continuous phenotypes, body mass index(BMI), cholesterol, high-density lipoprotein (HDL), low-density lipoprotein(LDL), from the UK Biobank(UKBB) dataset^45^. We then evaluated its performance internally on separate held-out UKBB testing datasets and externally on the independent All of Us (AoU) dataset. Results from these analyses demonstrate that DEEN achieved superior predictive performance for all phenotypes compared to existing methods. Moreover, the DEEN model significantly improved the stratification of various risk groups, demonstrating its potential for clinical utility.

## Results

### Overview of the DEEN algorithm

The DEEN algorithm comprises of three main components (Figure 1). Unsupervised autoencoder for learning latent representations: In the initial stage, DEEN employs an unsupervised autoencoder to derive lower-dimensional latent representations of the input SNPs for each chromosome. This approach leverages the expected correlations among the SNPs, such as those arising from linkage disequilibrium, to generate features that encode the independent variations among the SNPs. Each chromosome is modeled separately due to the minimal correlations expected across chromosomes. Concatenation of Latent Representations: In the second stage, the latent representations obtained from each chromosome are concatenated to form a combined feature set for predictive modeling. Supervised learning using an FCNN: The final stage involves using a supervised FCNN to model the chromosome-specific autoencoders. The FCNN can capture both linear and non-linear relationships among the input features, thus offering a broadened search space for identifying the optimal model. Notably, the FCNN inherently allows differential impacts of various genomic regions on the final prediction, thereby providing a more accurate representation of the underlying genetic effects. To evaluate the model, we employed 5-fold cross-validation on the UKBB dataset, splitting the data into 80% for training and 20% for testing. Within each training fold, we set aside 10% of the data specifically for hyperparameter optimization. Importantly, no test data was used in the optimization process. The trained models were subsequently evaluated for their predictive performance, risk stratification, and interpretation using the independent AoU dataset.

**Figure 1.**
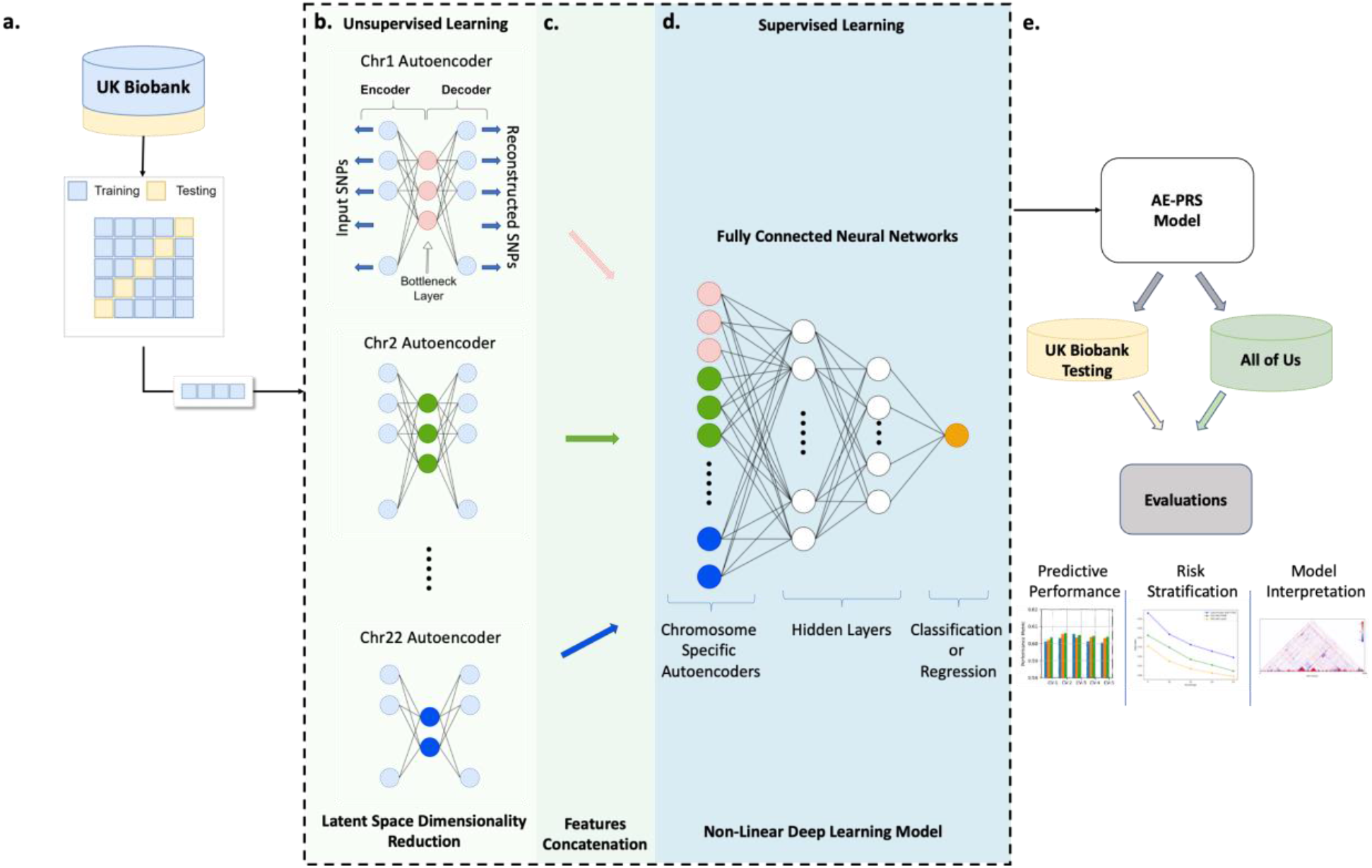
Overview of the study design and the DEEN algorithm. **a.** The UKBB dataset is divided into 5 equal parts for each phenotype. In each iteration, one-fold is used as the test set, while the remaining 4 folds are used for training. This process is repeated 5 times, with each fold used exactly once as the test set. **b**. Unsupervised latent space dimensionality reduction is performed using autoencoders with only genetic data. **c**. A single representation matrix for each patient is created by concatenating the latent space matrices obtained from the autoencoders. **d**. Supervised classification/regression with FCNN is carried out using the representation matrices obtained in part c and the phenotype data. **e**. Performance evaluations are conducted on the UKBB and All of Us datasets.

### Internal Evaluations of DEEN in UK Biobank

We optimized DEEN on UKBB training data for two binary phenotypes—hypertension and T2D as well as four continuous phenotypes: BMI, cholesterol, LDL, and HDL. The models’ predictive performance was assessed on the held-out UKBB testing data. We also evaluated existing PRS methods, including the GWAS summary-statistics based Plink_PT^46^, PRSice^15^, PRSCS ^26^ and Lasso (as implemented in bigsnpr) using individual-level data^47^, and an alternative ML method, PCA-FCNN, on the same datasets.

Figure 2a shows that DEEN achieved higher Area Under the Curve(AUC) scores for the two binary diseases compared to other evaluated PRS methods. In the UKBB dataset, DEEN’s AUC for T2D outperformed PLINK_PT by 6.27%, Lasso by 4.08%, PRSice by 3.39%, by 1.28%, and PCA-FCNN by 2.47%. For hypertension, DEEN showed similar gains: 6.15% over PLINK_PT, 3.21% over Lasso, 3.30% over PRSice, 2.31% over PRSCS, and 1.83% over PCA-FCNN.

**Figure 2.**
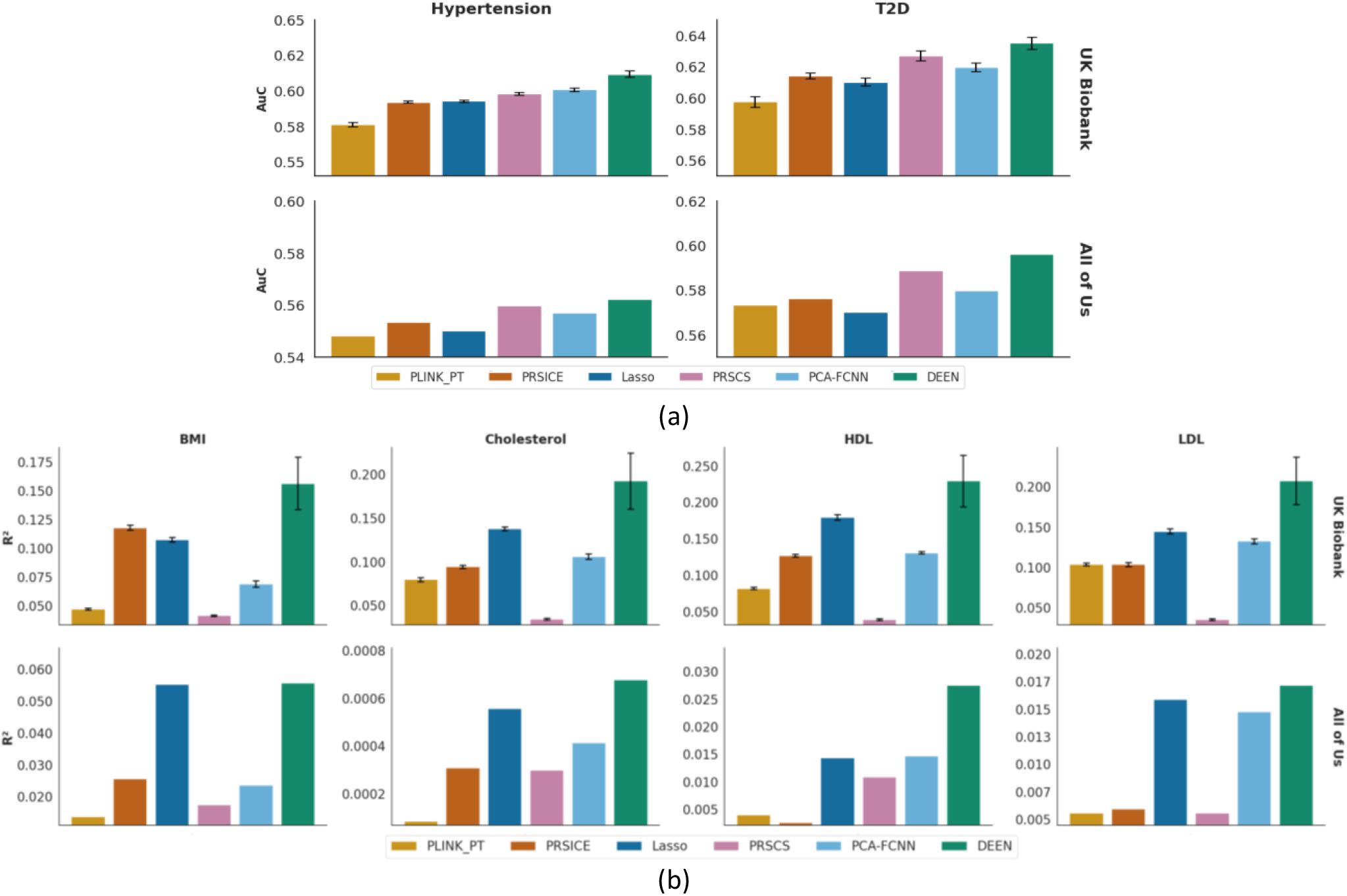
Predictive performances of DEEN and existing PRS methods on binary diseases and continuous traits in UKBB and AoU. The models compared in the figures are DEEN (green), PCA-FCNN (light blue), PRSCS (purple), Lasso (dark blue), PRSice(orange), and PLINK+PT (light orange). Error bars indicate the standard deviation of evaluation metrics across five testing folds for the UKBB dataset, reflecting variability in model performance. **a.** AUC values for different prediction models applied to Hypertension and T2D using data from the UKBB and AoU datasets. The top row shows the results for the UKBB dataset, with Hypertension on the left and T2D on the right. The bottom row shows the results for the AoU dataset in the same order. **b**. R^2^values of the different prediction models for continuous phenotypes: BMI, Cholesterol, HDL, and LDL phenotypes, respectively. The top row shows the results for the UKBB dataset, and in the bottom row, the results for the AoU dataset are shown in the same order as the given phenotypes.

Beyond predictive performance, we also evaluated the models’ ability to stratify risk for binary diseases, aligning more closely with their clinical utility. The analysis was performed by comparing the odds ratio enrichment of cases between high-risk and low-risk groups. Different risk quantiles (top 5%, 10%, 15%, 20%, and 25%) were used to select the high-risk group while the low-risk group was kept constant (bottom 5%). DEEN outperformed existing PRS approaches and PCA-FCNN in stratifying the two risk groups(Figure 3). For T2D, DEEN improved the odds ratio enrichment by an average of 58.27% compared to PLINK_PT, 25.12% compared to PRSice, 34.88% compared to Lasso, 8.66% compared to PRSCS, and 34.88% compared to PCA-FCNN. For hypertension, the respective average improvements are 84.87%, 37.12%, 42.30%, 31.70% and 3.74%.

**Figure 3.**
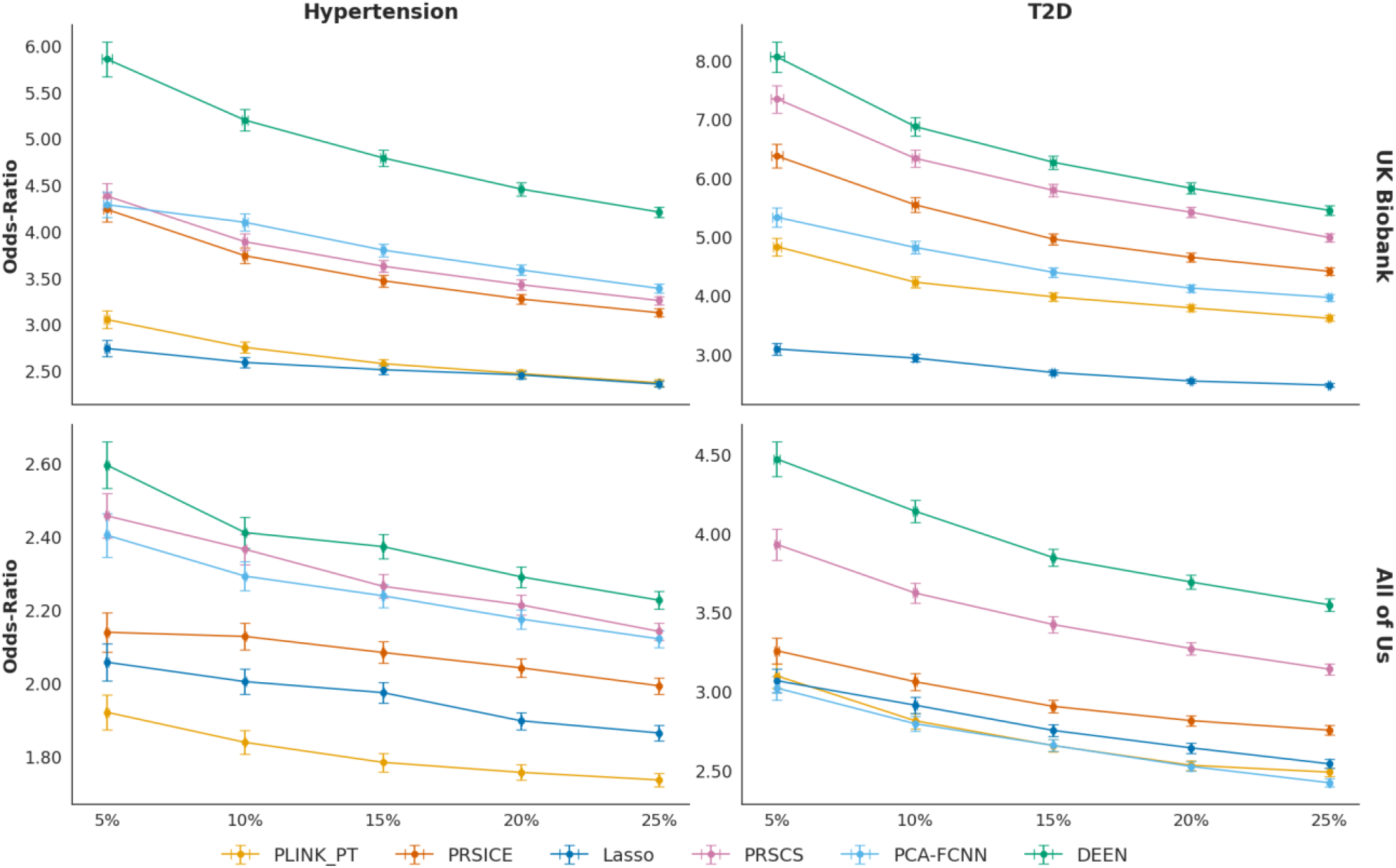
DEEN has improved risk stratification for T2D and Hypertension for the best-performing models. The models compared are DEEN (green), PCA-FCNN (light blue), PRSCS (purple), Lasso (dark blue), PRSice(orange), and PLINK+PT (light orange). The x-axis represents different percentage thresholds (5%, 10%, 15%, 20%, 25%) for selecting the high-risk group, and the y-axis shows the odds ratios enrichment of cases between the high- and low-risk groups for each threshold Error bars represent the 95% confidence intervals for the odds ratios. Results for UKBB are displayed in the top row, and for AoU are on the bottom row.

Figure 2b shows the performance evaluation of the models for continuous phenotypes. Consistent with the binary disease results, DEEN achieved higher *R*^2^ for all continuous phenotypes. For BMI, DEEN increased the *R*^2^ by 275.8% compared to PRSCS, 230.9% compared to PLINK_PT, 32.6 % compared to PRSice, 45.5% compared to Lasso and 126.2% compared to PCA-FCNN. For cholesterol, the *R*^2^ increase is 465.2%, 142.3%, 104.4%, 39.6%, and 81.8%, respectively. HDL showed the largest *R*^2^ increase, with improvements of 484.6%, 180%, 81%, 28%, 75.5%, and 169.8%. Lastly, for LDL, the *R*^2^ improvement is 489.6 %, 100%,100.6%, 43.3%, 56.7% and 158%, respectively.

### External Evaluations of DEEN in All of Us

External validation on the AoU dataset is shown in the bottom panels of Figure 2 and Figure 3. While the prediction AUC decreased on average for binary phenotypes across all methods, the proposed DEEN procedure still achieved the best results for all phenotypes. In addition, the odds-ratio analysis demonstrated that the improvement in stratifying risk groups remained stable. In each case, the DEEN method outperforms the other methods.

For continuous phenotypes, the DEEN model applied to the AoU dataset continued to outperform other methods, similar to the results obtained with the UKBB dataset. In contrast, the performance of other methods varied. Lasso outperformed PRSice for BMI, while PRSCS outperformed PRSice for Cholesterol and PRSice for HDL.

### DEEN Transformed Latent Features Capture Local and Distal Genetic Relationships

To further investigate the improved performance of DEEN compared to the leading existing PRS methods, the contributions of SNPs to the final predictions were compared using model outputs from the T2D analysis (Figure 4). As DEEN separates dimensionality reduction and predictive modeling, most SNPs were retained by the model as they may contribute to either reconstructing the genetic features or predicting the outcomes. In contrast, approximately 90% of the SNPs have coefficients of zero in the lasso model or values close to zero in PRSCS, as these models only retain SNPs that are both strongly correlated with the outcome and representative of the feature space. Therefore, DEEN comparatively utilizes more SNPs when constructing the prediction models. Similar results were observed for other diseases (Supplementary Figure S7-S11).

**Figure 4.**
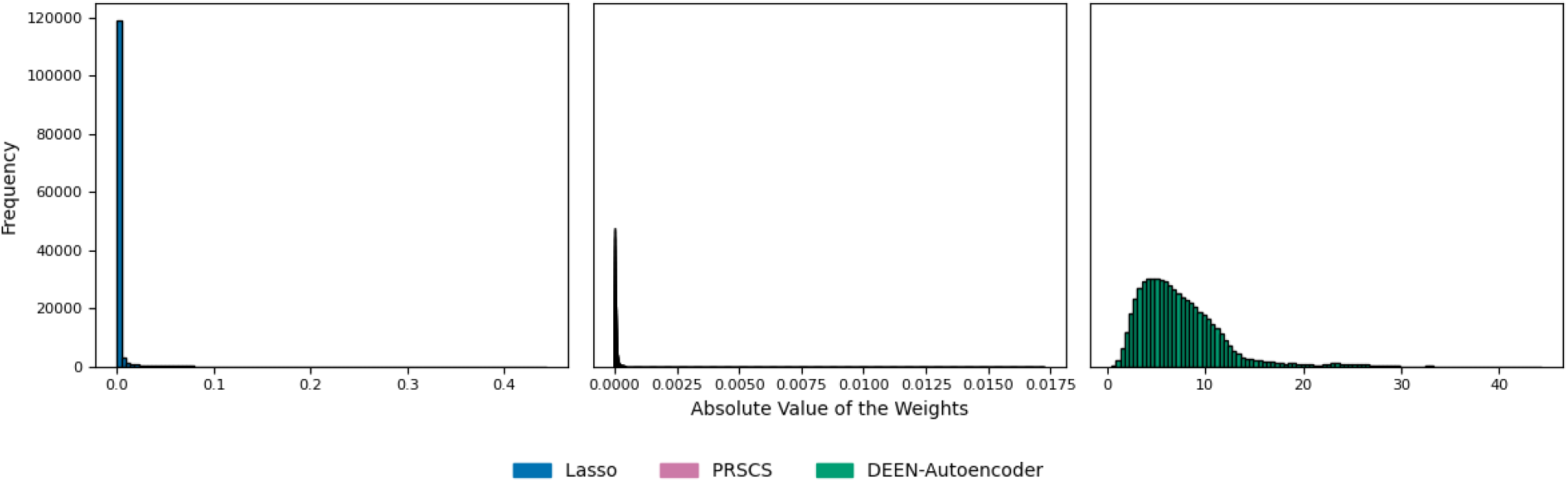
Comparison of SNP contributions between autoencoder, PRSCS, and Lasso. Each histogram shows the frequency distribution on all chromosomes of the respective SNP weight values of the Lasso regression (left), PRSCS (center), and autoencoder model (right) for T2D disease.

The autoencoder can also capture both local and distal relationships among the SNPs. Using autoencoder outputs from the same T2D analysis, Figure 5a shows the Pearson correlations among the SNPs based on their estimated feature weights in the autoencoder model. Since the SNPs are organized according to their physical locations across the chromosomes, SNPs in proximal distances showed the highest correlations, reflecting local linkage disequilibrium. However, the autoencoder model also captured correlations among SNPs in distant regions, which are often not modeled by existing PRS methods.

**Figure 5.**
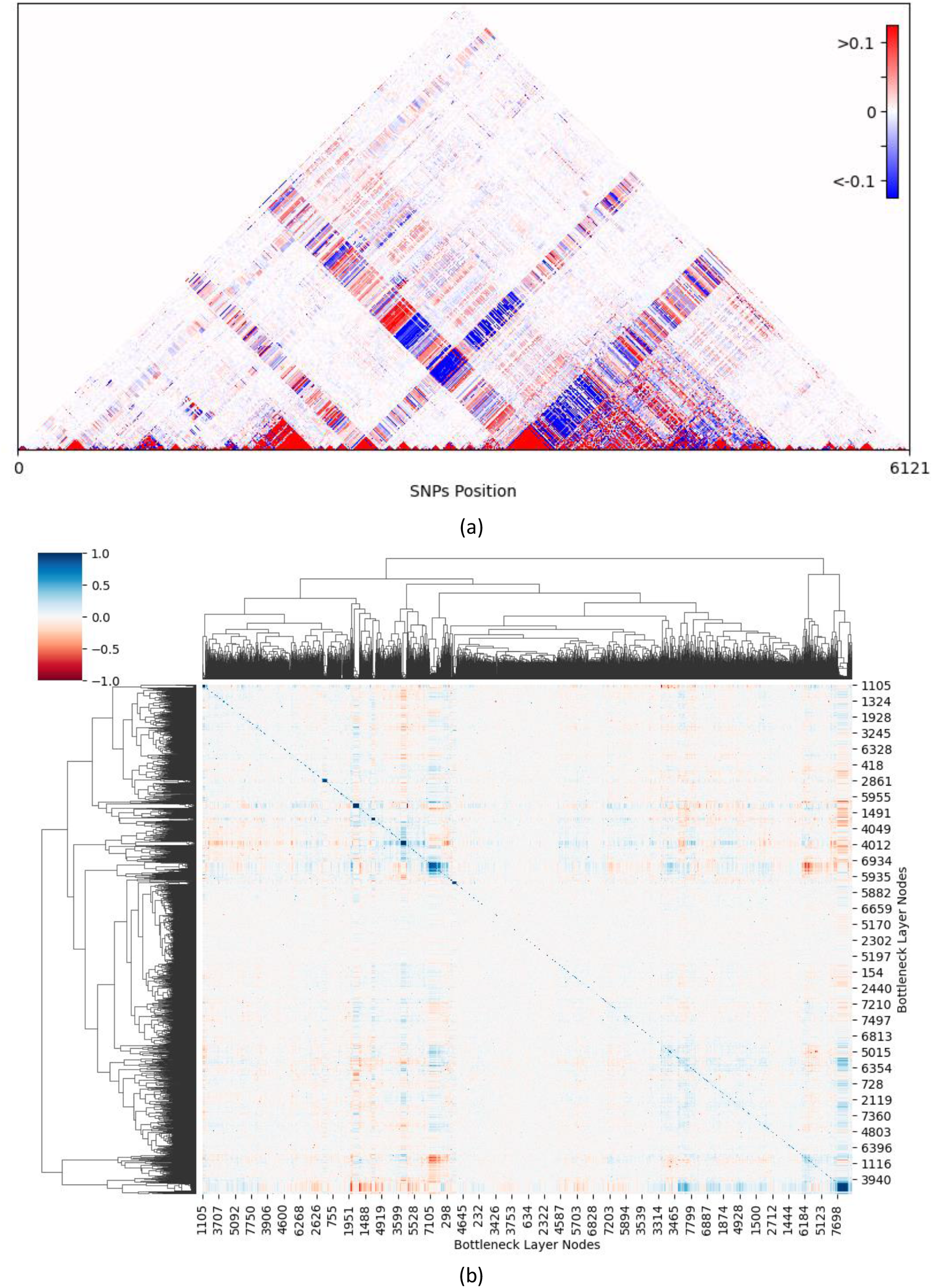
Correlations in the Encoder Layer Nodes Weights and Cluster Map of Correlations in the Decoder Layer Nodes Weights. Autoencoder output from chromosome 1 in the T2D analysis is used to demonstrate the SNPs and node correlations. **a.** correlations of the weights of the encoder SNP inputs, calculated using the Pearson correlation coefficient. Red indicates positive correlations, while blue indicates negative correlations. **b**. cluster map showing the correlation values between nodes in the decoder layer of chromosome specific autoencoder. The heatmap visualizes correlations ranging from -1 to 1 representing positive correlations in red and negative correlations in blue, while white indicates no correlation.

Additionally, the nodes in the bottleneck layer can differentially model the SNPs. Comparing the SNPs’ weights connected to each node in the bottleneck layer revealed distinct groups of nodes with similar SNP connections, indicating that the autoencoder has distinct structures modeling different parts of the genome (Figure 5b). Results for other diseases are presented in the Supplemental Materials (Figure S18-S23).

## Discussion

In this study, we introduce a novel method for computing PRS using deep learning, employing a deep ensemble encoder-based approach that integrates autoencoders and FCNNs. DEEN differentiates itself from existing PRS methods by modeling non-linear genetic effects through more flexible structures. This allows SNPs in different genomic regions to exert differential impacts on the final prediction via learned weights, unlike traditional PRS methods that apply uniform priors or regularization penalties to all SNPs. Furthermore, DEEN addresses the common issue of overfitting in deep learning models by initially employing an autoencoder model to learn a reduced latent dimension of the genetic data. Given the complexity and size of genetic datasets, directly applying a deep learning model is impractical due to computational constraints such as GPU memory limits. DEEN overcomes this by separating dimensionality reduction and final predictive modeling into distinct modules. The autoencoder module significantly reduces the correlation among input SNPs, such as those driven by LD, creating a new latent representation. This reduced input feature space allows the FCNN to build more efficient and effective predictive models of disease.

Through real data analysis in the UKBB, we demonstrated that DEEN consistently demonstrates superior predictive performance for both binary diseases and continuous traits compared to existing PRS methods, including PLINK_PT, PRSice, PRSCS, Lasso regression, and an alternative ML method, PCA-FCNN (Figure 2). In addition, risk stratification analysis, which aligns more closely with the clinical utility of risk scores, showed that the DEEN generated risk scores significantly improved our ability to identify individuals in the high-risk group (Figure 3). Since individuals in the high-risk group are more likely to benefit from preventive schemes or treatment options, the proposed model is more likely to provide clinically relevant value. We also independently validated the DEEN models in the AoU dataset. Although slight decreases in performance were observed due to population, demographic, and environmental differences between biobanks in different countries, the predictive performance remained higher than that of other methods.

We showed that the disentanglement of dimensionality reduction of genetic data and predictive modeling of the outcome improved the predictability of DEEN. Both PCA-FCNN and DEEN, which perform separate dimensionality reduction and predictive modeling, have shown improved performance compared to existing PRS methods (Figure 2). On the other hand, autoencoder has been shown to be highly efficient in extracting lower dimensions of data. Compared to dimensionality reduction using principal component analysis (PCA), the autoencoder model has learned a better data representation, as evidenced by the improved final prediction using the same FCNN model as the prediction module (Figures 2 and 3). When we compared the model weights of the same SNPs from the autoencoder model, Lasso and PRSCS PRS, it was clear that the former model encompasses more SNPs (Figure 4). SNPs included in the DEEN model may be informative in either learning the latent representations or predicting the outcome, or both. In contrast, in Lasso and PRSCS PRS, SNPs that are not predictive of the outcome are removed from the model, even if they may be important in modeling the correlations among SNPs. Finally, the autoencoder can capture both proximal and distal SNP relationships (Figure 5a). These relationships are modeled through the bottleneck layers of the autoencoder, which has shown distinct clusters among the nodes (Figure 5b). The clustering among the bottleneck nodes indicates that different parts of the network are differentially modeling groups of SNPs. These results demonstrate that DEEN is more flexible in modeling genetic data in relation to predicting outcomes compared to existing PRS methods.

The study also has several limitations that warrant future research. The DEEN method requires individual-level genetic and phenotype data to train and optimize the model. On the other hand, the Plink_PT, PRSice and PRSCS methods only require GWAS summary statistics to generate the PRS. As a result, DEEN is more computationally expensive than summary statistics-based PRS methods, but with a gain in predictive performance. In addition, to reduce potential model overfitting and stay within the computational constraint (e.g. GPU memory), we performed the necessary variant filtering based on existing GWAS results. While the variant filtering may remove potential SNPs with small effects, the filtering was consistently applied to all methods to ensure valid comparisons. Furthermore, we evaluated multiple filtering thresholds, and the relative performances among different methods were stable. Lastly, the DEEN model may be less interpretable than PRS generated from statistical models due to the complexity of deep learning models. Developing interpretable machine learning models could potentially be incorporated into DEEN in the future.

Future research could investigate the potential of autoencoders as transfer learning tools to improve model performance across diverse racial groups. Additionally, prioritizing efforts to improve the interpretability of autoencoder models without sacrificing performance, optimizing computational efficiency during training, and expanding their applications to other complex diseases or datasets are crucial steps forward. Incorporating additional external validation datasets can further strengthen the generalizability of findings beyond those utilized in this study.

In conclusion, to our knowledge, our study is the first to demonstrate the benefits of ensembling advanced machine learning algorithms, autoencoder and FCNN, for generating improved PRS. Our study provides evidence that modeling complex genetic effects can improve the genetic risk prediction of complex diseases and traits. We also have made the DEEN algorithm publicly available on GitHub for replications and evaluations.

## Material and methods

The UKBB Dataset is a comprehensive biomedical database supporting health research in the United Kingdom and worldwide^48–51^. The data collected from more than 500,000 volunteers aged 40-69 living in the UK includes health questionnaires, electronic health records (EHR), physical measurements, biological samples, genetic information, imaging data, and digital health data. The AoU Dataset, which is used as external validation in this study, is a dataset containing comprehensive genetic data collected within the scope of the AoU Research Program conducted by the US National Institutes of Health (NIH)^52^. This dataset contains the genetic data of more than 200,000 participants of different races.

### Data quality control and variants selection

We performed a series of quality control (QC) measures to ensure the quality of the analyzed dataset. For sample-level QC, we retained individuals who passed the following criteria: (1). Only unrelated individuals were retained. Among related individuals, one individual from each pair was systematically removed to prevent undue influence from familial genetic connections. The threshold for relatedness was set at the level of second-degree relatives, as indicated by an identity-by-descent ^π value equal to or greater than 0.25. (2) Individuals with self-reported White British ancestry. This ensures compatibility with the population used to generate pre-trained PRS. (3) Individuals with matched self-reported and genetically inferred sexes. (4). Individuals with heterozygosity within three standard deviations from the mean. For SNP-level QC, we excluded SNPs that: (1) with more than 5% missing rate, (2) minor allele frequency of less than 1%, (3) have an imputation quality info score of less than 0.8 (4) are duplicated or ambiguous (5) Hardy-Weinberg equilibrium p-value less than 10^−10^.

For hypertension and T2D, case and control statuses were determined using the widely adopted PheCode algorithm in UKBB, which relies on the inclusion and exclusion of disease-specific ICD-10 codes^53,54^. The continuous traits were extracted from the UKBB data fields: field 21001 for BMI, field 30690 for cholesterol, field 30760 for HDL, and field 30780 for LDL. The study cohort in the AoU dataset was selected to consist of individuals who self-identified as White, as indicated by the data in column 1586140. This selection was made to ensure that the ancestry of these individuals is consistent with that of the participants in the UKBB, facilitating a more comparable analysis between the two datasets. The corresponding phenotyping algorithms for binary diseases were implemented by the AoU team^55^. The corresponding continuous traits were obtained from AoU with concept IDs 3038553 for BMI, 40772590 for cholesterol, 40782589 for HDL, and 40795800 for LDL.

We further selected individuals whose BMI, Cholesterol, HDL, and LDL values fell within three standard deviations of the mean to exclude extreme outliers. To address potential distributional differences between UKBB and AoU, we applied the same filtering criteria to AoU using thresholds defined by the UKBB dataset. Specifically, trait values were filtered as the following: BMI from 13.136 to 41.679 kg/m^2^, cholesterol from 41.022 to 164.7 mg/dL, HDL from 5.49 to 46.782 mg/dL, and LDL from 17.244 to 111.348 mg/dL. Additional details regarding the train-test split and case-control counts for binary phenotypes are provided in Table 1, while Table 2 summarizes the train-test counts for continuous traits.

**Table 1.** Sample sizes for binary phenotypes for UKBB and AoU datasets.

| Phenotype | UKBB Dataset |  |  |  | AoU Dataset |  |
| --- | --- | --- | --- | --- | --- | --- |
|  | Training |  | Testing |  | Testing |  |
|  | Case | Control | Case | Control | Case | Control |
| **Hypertension** | 94,985 | 222,767 | 23,747 | 55,692 | 40,791 | 82,624 |
| **T2D** | 24,747 | 291,206 | 6,147 | 72,802 | 17,737 | 105,678 |

**Table 2.** Sample sizes for continuous traits for UKBB and AoU datasets.

| Phenotype | UKBB Dataset |  | AoU Dataset |
| --- | --- | --- | --- |
|  | Training | Testing |  |
| **BMI** | 316,878 | 79,220 | 120,524 |
| **Cholesterol** | 303,076 | 75,769 | 60,908 |
| **HDL** | 277,357 | 69,340 | 56,121 |
| **LDL** | 302,521 | 75,631 | 58,196 |

In addition, when applying the proposed DEEN method, we performed variant filtering to reduce computational time and potential null signals. For each binary trait, we selected candidate SNPs associated with the trait using a p-value threshold of less than 0.05, 0.005, or 0.0005 based on GWAS results in the UKBB training dataset. The respective GWAS was conducted using PLINK 2.0, adjusting for age, gender, ancestry principal components, and assessment center as covariates. For continuous traits, 100,000 or 150,000 SNPs with the lowest p-values were selected so, ensuring the number of SNPs was comparable to the binary diseases^56^. The entire analysis was repeated for all thresholds.

### Polygenic risk score methods

The PRSCS method is a Bayesian approach used to more accurately predict the effects of genetic variants on a set of diseases or phenotypes^26^. For our analysis, we used the default parameters of PRSCS along with automatic tuning of global shrinkage parameter phi and n_gwas=320000, which specifies the sample size of the GWAS. For the LD reference genome, we used the precomputed LD reference using the UKBBdata provided by the software. The GWAS summary statistics were obtained from the published GWAS summary statistics of the UKBB^56^. The models were fitted with p-value selected or HAPMAP3 SNPs per package’s recommendation. Lasso (Least Absolute Shrinkage and Selection Operator) is a regression method used for variable selection and regularization, especially for high-dimensional data. We used the Lasso method implemented in the BigSNPr^47^ package to generate the Lasso PRS used in this study. Specifically, we used the big_spLogReg and big_spLinReg functions for binary and continuous phenotypes, respectively. The lambda tuning was conducted using internal cross-validation with the k parameter set to 5, representing the number of folds used in cross-validation. These parameter settings were selected using the default parameters provided by the respective methods.

PRSice is a software tool used for analyzing PRS. It is designed to efficiently calculate and summarize risk scores based on GWAS summary statistics and individual genotype data. PRSice performs clumping of SNPS at r^2^ = 0.1. We set P value thresholds of input SNPs at P < 0.05, 0.005, and 0.0005 for different experiments. We applied PRSice with the following parameters: interval=0.0005, score=sum, model=add.

We also utilized the PLINK_PT method, which applies a pruning and thresholding (P+T) approach to select independent and statistically significant genetic variants. This method ensures that the SNPs included in the PRS calculation are minimally correlated and meet specific p-value thresholds, improving the robustness of the resulting PRS. We also used --score function from PLINK2 to calculate PRS. For each individual j, the PRS is calculated by summing the contributions from each genetic variant. The contribution of each variant is determined by two factors: βi, which is the weight for each variant based on its log odds ratio, and Gij, the number of risk alleles (0, 1, or 2) of variant *i* present in individual *j*. In other words, the PRS for an individual is the total of these weighted values for all genetic variants considered.

To ensure fairness, all methods used the same input SNPs for comparative analyses.

### Dimensionality Reduction with PCA

As a comparison to our proposed method, we also utilized a common method for dimension reduction, the principal component analysis^57^ method, implemented in Sklearn^58^ v1.3.1. For each chromosome, we applied PCA to reduce the dimensionality of the genetic feature space. We evaluated the PC space between 5% and 50% of the original dimensions. For each PC dimension, we calculated the variance explained and for each chromosome, we calculated the number of PCs required for the variance to exceed 90%.

The number of principal components (PCs) required varied across chromosomes. To address this variation, we determined the number of PCs proportionally to the number of SNPs in each chromosome. Specifically, the number of PCs was calculated as the total SNP count divided by 8, ensuring that the variance explained ranged between 93-97% for all chromosomes. This method preserves genetic diversity while effectively reducing dimensionality. For instance, chromosome 1, associated with hypertension and containing 55,815 SNPs, was proportionally reduced to 6,976 nodes in the lower-dimensional space. Similarly, chromosome 22, which includes 9,480 SNPs, was reduced to 1,180 nodes. This proportional scaling ensures that chromosomes with a higher SNP count retain sufficient dimensionality, while smaller chromosomes are adequately represented in the reduced feature space.

The dimension-reduced patient array for this method can be given as follows:

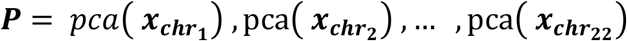

where **x** is the input array of the chromosome. The computed patient array is then given as input to the FCNN. The modeling process of combining PCA and FCNN is referred to as PCA-FCNN in this study.

### Dimensionality Reduction with Autoencoders

The proposed DEEN method consists of 3 main parts, as shown in Figure 1. In the first part, autoencoders were used for dimensionality reduction. In this study, PyTorch^59^ and PyTorch Lightning^60^ libraries were used to train and evaluate autoencoder and FCNN models. An autoencoder is a type of artificial neural network designed to learn efficient coding of input data through compression. It primarily consists of two components: an encoder that maps the input data to a latent space representation and a decoder that reconstructs the input data from this compressed representation. During this process, the model learns the key features of the original input. A learning process takes place to ensure that the compressed representation is as similar as possible to the original data. In this study, grid search was used for hyperparameter selection to optimize the performance of the model. Each of the hyperparameters is tested individually by trials to determine the optimal values. This manual grid search process allowed for a detailed analysis of the impact of each hyperparameter on the performance of the model and allowed for more fine-tuning.

Separate autoencoders were trained for each chromosome using the training dataset. The number of nodes in the bottleneck layer required for each chromosome was kept the same as the number of dimensions determined by the PCA experiments. The coded features of the patients in the training and test dataset were obtained using only the coder blocks of the autoencoders obtained after training. The set of hyperparameters required for training the autoencoders was the number of layers, batch size, learning rate, weight decay, activation functions, and epoch size. Mean Squared Error(MSE) was used as the loss function during autoencoder training. The parameters used in the experiments for the grid search are learning rate 0.0001, 0.00001, and 0.000001, weight decay 0, 0.001, and 0.1, epoch size 100,200,400, chunk size 64,256,1024 and the number of layers 2,3 and 4. Through gird search optimization, the following parameters were determined: learning rate 0.00001, weight decay 0, epoch size 400, batch size 256, and number of layers 2. The same parameters were used for all diseases. The activation function used between the layers is ReLU^61^. Details of these experiments can be found on the GitHub page https://github.com/Cedars-CIG/DEEN.

The encoded chromosome is represented by a function where k is the bottleneck layer,

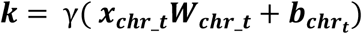

where ***x***_***chr***_***k***_ is the input matrix of the chromosome *t*, ***W***_***chr***_***k***_ is the matrix of weights between the input and encoder layer for chromosome *t*, b is the vector of biases for the encoder layer, and γ : ℝ → ℝ an activation function. Similarly, the decoding network can be formulated as follows,

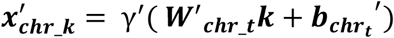

where ***W***^′^_***chr***_***t***_ is the weight matrix between the encoder and output layer of the autoencoder. The loss function for each chromosome is given as:

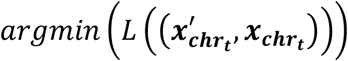

where L is the loss function. We can write the resulting loss function with MSE as follows:

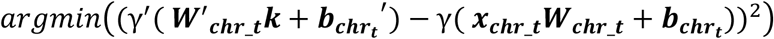

After this minimization process, Patient array P is calculated using the calculated weights as follows:

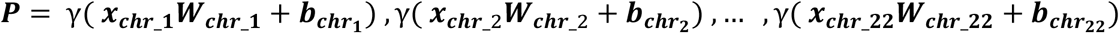

The computed patient array is then given as input to the FCNN.

### Fully Connected Neural Networks

FCNN is a widely used model in artificial neural networks. FCNN consists of layers where each neuron is connected to all neurons in the previous layer. These networks usually operate as feedforward networks, meaning that information flows unidirectionally from the input layer of the network to the output layer. After dimensionality reduction in both PCA-FCNN and autoencoder-based methods, the reduced data was used for classification or regression with FCNN. The hyperparameter optimization for this method was similar to the autoencoder method. The set of hyperparameters required to train the FCNN is the number of layers, node size, batch size, learning rate, weight decay, activation functions, and epoch size. These hyperparameters were determined by conducting separate training for each phenotype. As with the autoencoder method, a manual grid search was performed on pre-defined values to obtain the best results. These experiments were carried out using several network models with 2,3, and 4 layers and sizes ranging from 16 to 2048 with learning rates 0.0001 and 0.00001, weight decay 0, 0.001, and 0.1, and batch sizes 256-512-1024.

This patient array will be used as input to the first hidden layer of the classification/regression network:

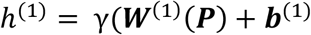

Between hidden layers:

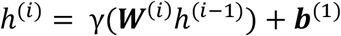

Between the last hidden layer to the classification layer:

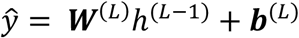

where L is the decision layer, and i is the layer number of the classification/regression network.

Binary Cross Entropy(CE) was used as the loss function in the classification models, while MSE was used in the regression models. The formulation of MSE and BCE were given as,

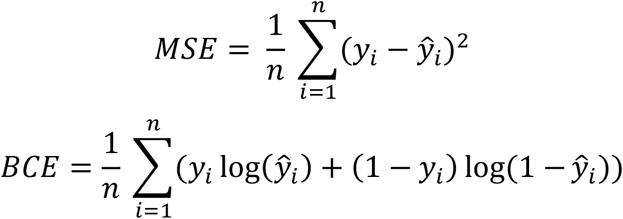

where *y*_*i*_ is the observed value of the patient *i, ŷ_i_* is the estimated value of the patient *i*, and n is the number of patients.

### Correlation Analysis with Autoencoders

Autoencoders can identify important patterns among SNPs and generate a lower-dimensional encoding (latent space) representation of these SNPs. During the learning process, each input SNP is connected to neurons in the hidden layer of the model through learned weights. These weights determine how the model processes the input data and highlight the importance of each SNP. By analyzing the correlation of these weights, we can identify associations between different input SNPs. E.g. a high correlation between the weights of two SNPs may suggest that they contribute similarly to the model. In this study, we examined the correlation between the weights of the autoencoders trained separately for each chromosome to investigate correlative relationships among SNPs. These relationships can indicate potential linear or non-linear interactions that are important for learning the latent representation. Conversely, analyzing correlations between nodes in the bottleneck layers can reveal nodes that differentially model groups of SNPs. Such correlations could indicate that the model captures the finer and more complex structure of the data.

## Method Evaluations

To thoroughly evaluate our model’s performance, we implemented a 5-fold cross-validation strategy using the UKBB dataset. The dataset was evenly divided into five equal subsets. In each iteration, one subset served as the test set, while the remaining four subsets formed the training set. This process was repeated five times, allowing every data point in the UKBB dataset to contribute to the evaluation.

For all models, including PRSCS, PRSice, PLINK, LASSO, and DEEN, we strictly ensured that the training data was used solely for training purposes. GWAS analyses were also conducted exclusively on the training data. Specifically for DEEN, both the autoencoder and classification/regression models were trained without any involvement of test data, thereby preserving the integrity of the evaluation. Hyperparameter optimization was performed within the training data of each fold, using a 90-10 split, with 90% allocated for training and 10% for validation. For LASSO, the internal cross-validation was conducted exclusively within the training data to tune the lambda parameter without incorporating test data.

After completing the training and validation process within the UKBB data through this cross-validation framework, the optimized models were applied to the AoU dataset. The AoU dataset was used solely for external evaluation and was entirely excluded from the training process. This independent evaluation provided a robust measure of the models’ generalizability and performance across different datasets.

The study used two performance metrics, R^2^ for continuous traits and AUC for binary diseases for evaluating the predictive performances of PRS models. R^2^ is a commonly used error measure in regression problems. It is a statistical measure that represents the proportion of the variance for a dependent variable that is explained by an independent variable or variables in a regression model. AUC evaluates the True Positive Rate (TPR) against the False Positive Rate (FPR) at various thresholds. TPR measures the proportion of actual positive cases that are correctly identified by the model, while FPR measures the proportion of actual negative cases that are incorrectly classified as positive by the model. AUC takes a value between 0 and 1, with a value closer to 1 indicating better classification ability of the model.

To assess the risk stratification of PRS models, we calculated the odds ratio of case enrichment between high-risk and low-risk individuals. We stratified individuals into high-risk and low-risk groups based on their predicted probabilities. For the low-risk group, we selected individuals with the lowest 5% predicted probabilities according to each model. Conversely, we varied the high-risk group selection for each model by progressively relaxing the predicted probability threshold from the top 5% to the top 25%, in increments of 5%. A logistic regression model was then employed to determine the odds ratio enrichment of cases between the high-risk and low-risk groups for each model at each threshold.

The development and training of deep learning models was carried out using the PyTorch and PyTorchLightning libraries. In the training process, we used a hardware configuration with a 32-core CPU, 1 NVIDIA A100 GPU, and 100 GB RAM.

## Data Availability

The UKBB is a large-scale biomedical database with genetic and health information from more than 500,000 UK participants. Available for research by request at https://www.ukbiobank.ac.uk. The UKBB data was approved under application # 86494 The All of Us Research Program is a large-scale biomedical database with diverse health data from over one million U.S. participants. Available for research by request at this link: https://www.researchallofus.org/

https://www.ukbiobank.ac.uk

## Data and Code availability

The UKBB is a large-scale biomedical database with genetic and health information from more than 500,000 UK participants. Available for research by request at https://www.ukbiobank.ac.uk. The UKBB data was approved under application # 86494 The All of Us Research Program is a large-scale biomedical database with diverse health data from over one million U.S. participants. Available for research by request at this link: https://www.researchallofus.org/ We provide the scripts used to perform the model training and inference proposed in the article in the GitHub repository https://github.com/Cedars-CIG/DEEN.

## Acknowledgments

“The All of Us Research Program is supported by the National Institutes of Health, Office of the Director: Regional Medical Centers: 1 OT2 OD026549; 1 OT2 OD026554; 1 OT2 OD026557; 1 OT2 OD026556; 1 OT2 OD026550; 1 OT2 OD 026552; 1 OT2 OD026553; 1 OT2 OD026548; 1 OT2 OD026551; 1 OT2 OD026555; IAA #: AOD 16037; Federally Qualified Health Centers: HHSN 263201600085U; Data and Research Center: 5 U2C OD023196; Biobank: 1 U24 OD023121; The Participant Center: U24 OD023176; Participant Technology Systems Center: 1 U24 OD023163; Communications and Engagement: 3 OT2 OD023205; 3 OT2 OD023206; and Community Partners: 1 OT2 OD025277; 3 OT2 OD025315; 1 OT2 OD025337; 1 OT2 OD025276. In addition, the All of Us Research Program would not be possible without the partnership of its participants.”

R.L is supported in part by Cedars Sinai Department of Neurology/Jona Goldrich Center for Alzheimer’s & Memory Disorders. O.B.O, R.C, and R.L are supported by the Department of Computational Biomedicine, Cedars Sinai.

## Author Contributions

O.B.O. and R.L. contributed to method development, model training, and writing the paper, while R.C. contributed to data processing.

