## Supplemental Material for "A Deep Ensemble Encoder Network Method for Improved Polygenic Risk Score Prediction" (from another version of this preprint: version 1, 2024-08-02)

a.
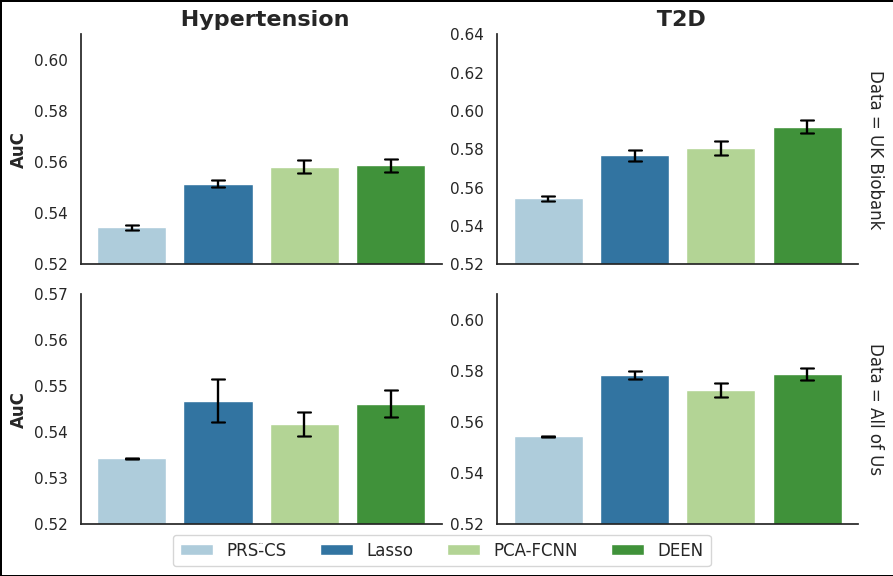

b.
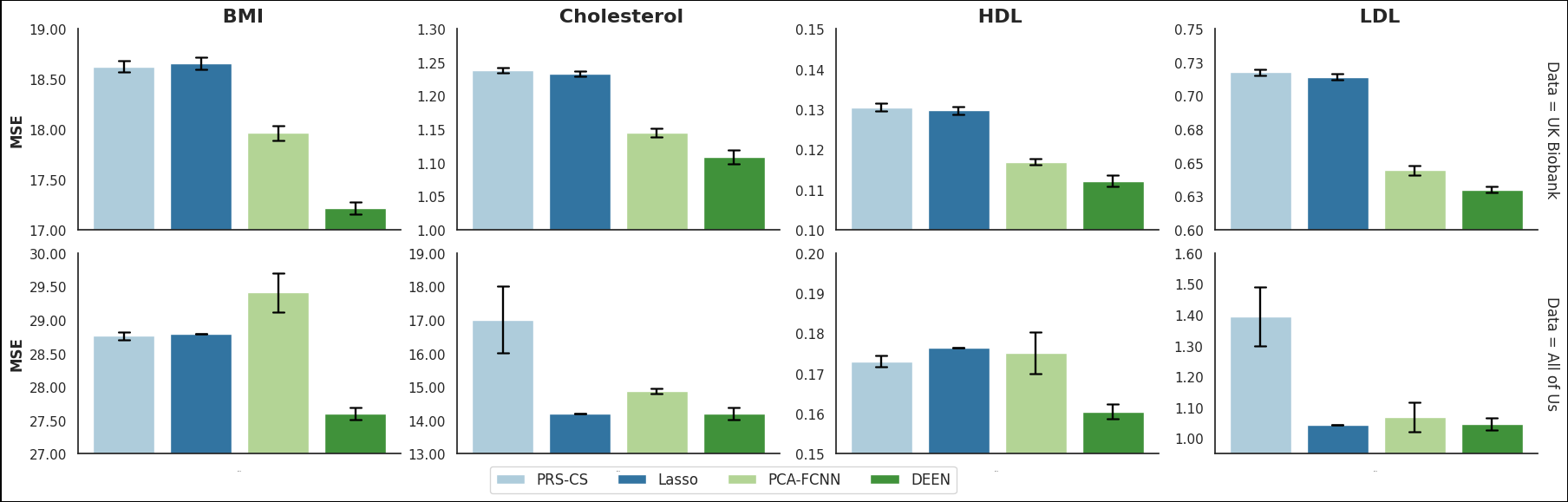

Figure S- 1 Predictive performances of DEEN and existing PRS methods on binary diseases and continuous traits in UKBB and AoU for P<0.0005. The models compared in the figures are DEEN (green), PCA-FCNN (light green), Lasso (dark blue) and PRS-CS (light blue). **a.** AuC values for different prediction models applied to Hypertension and T2D using data from the UK Biobank and All of Us datasets. The top row shows the results for the UK Biobank dataset, with Hypertension on the left and T2D on the right. The bottom row shows the results for the All of Us dataset in the same order. **b.** MSE values of the different prediction models for continuous phenotypes: BMI, Cholesterol, HDL, and LDL phenotypes respectively. The top row shows the results for the UK Biobank dataset and in the bottom row, the results for the All of Us dataset are shown in the same order as the given phenotypes.

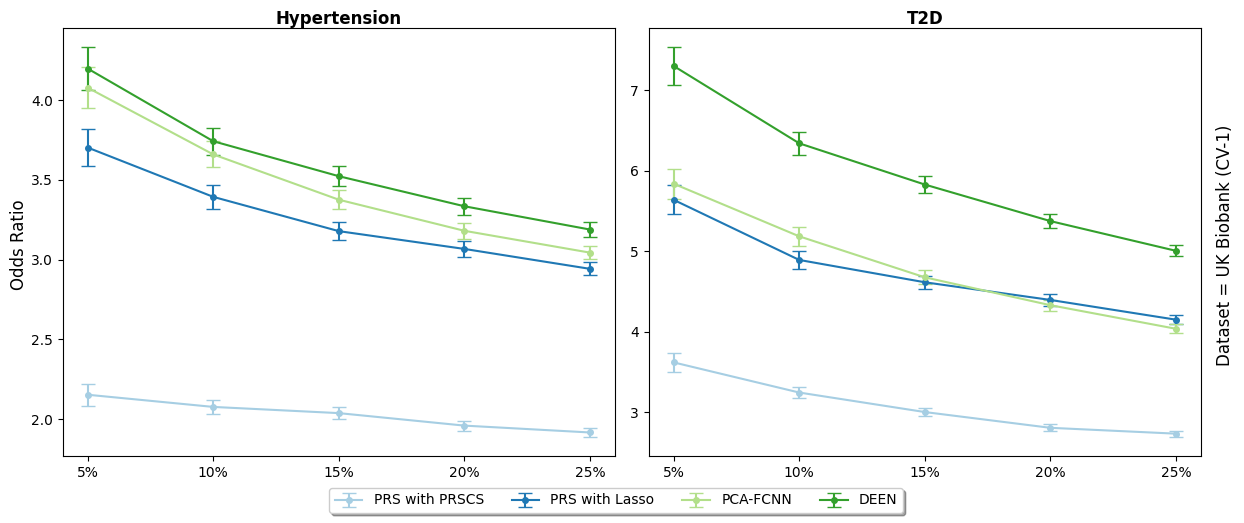

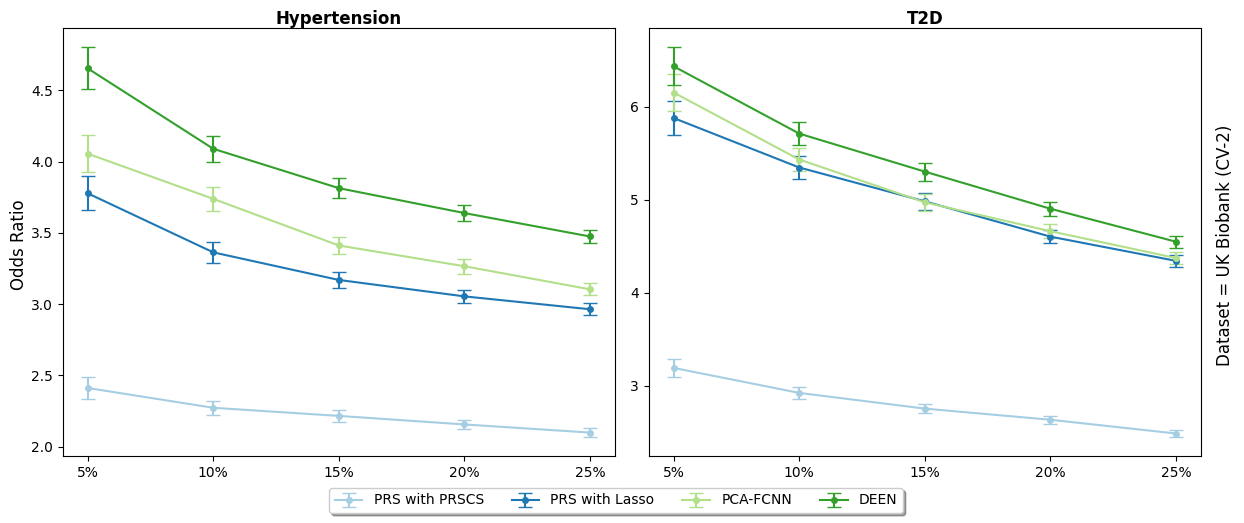

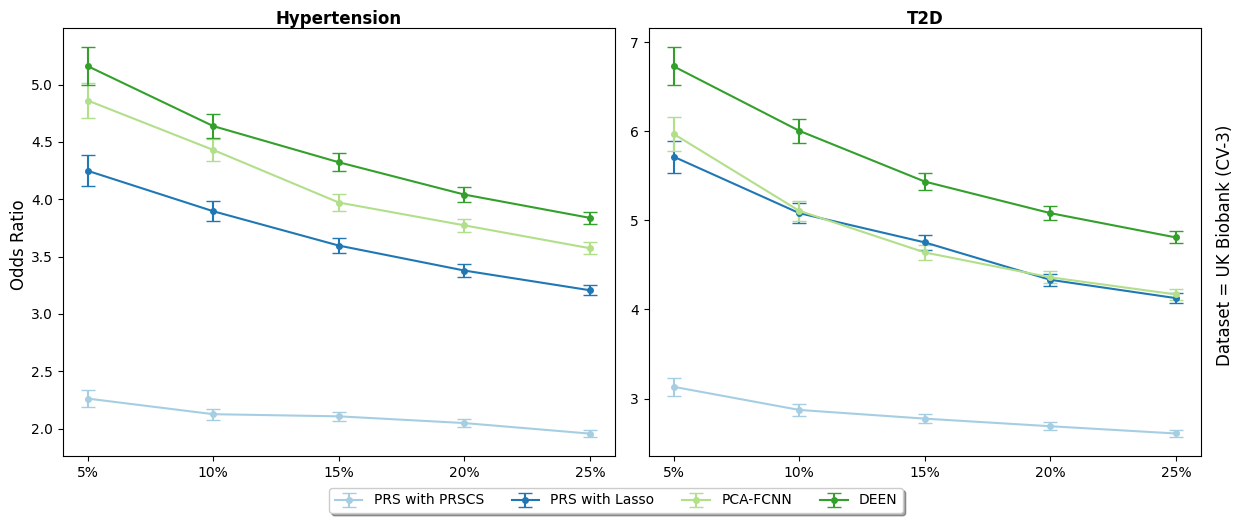

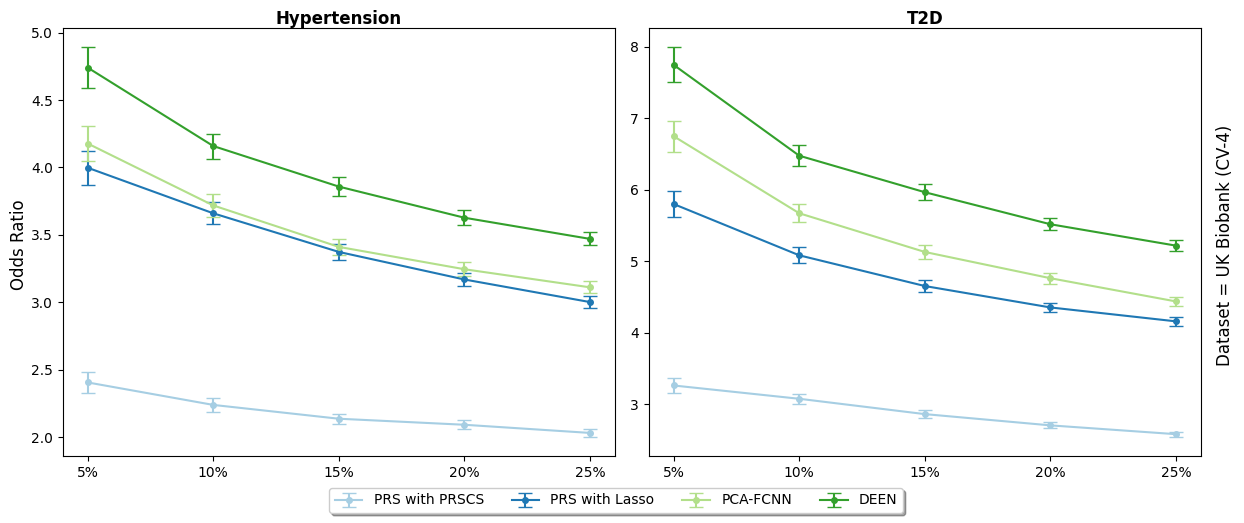

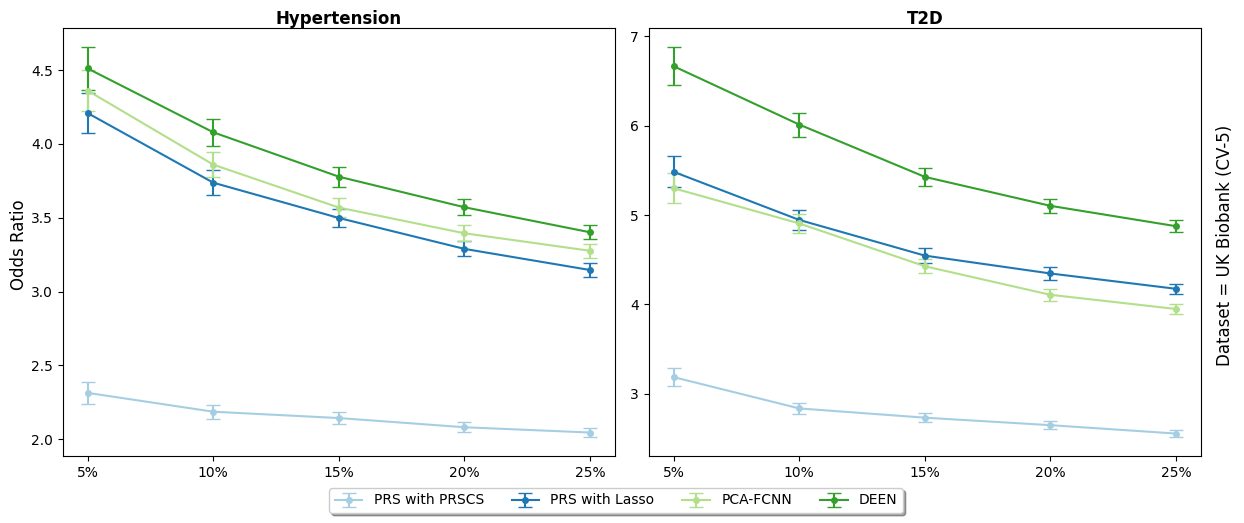

Figure S- 2 DEEN improved risk stratification for T2D and Hypertension Phenotypes for UKBB with p<0.005 for CV-2 to CV-5. The models compared are DEEN (green), PCA-FCNN (light green), Lasso (dark blue), and PRSCS (light blue). The x-axis represents different percentage thresholds (5%, 10%, 15%, 20%, 25%) for selecting the high-risk group, and the y-axis shows the odds ratios enrichment of cases between the high- and low-risk groups for each threshold. 95% confidence intervals are shown for each odds ratio. Results for UKBB are displayed in the top row, and for AoU are on the bottom row.

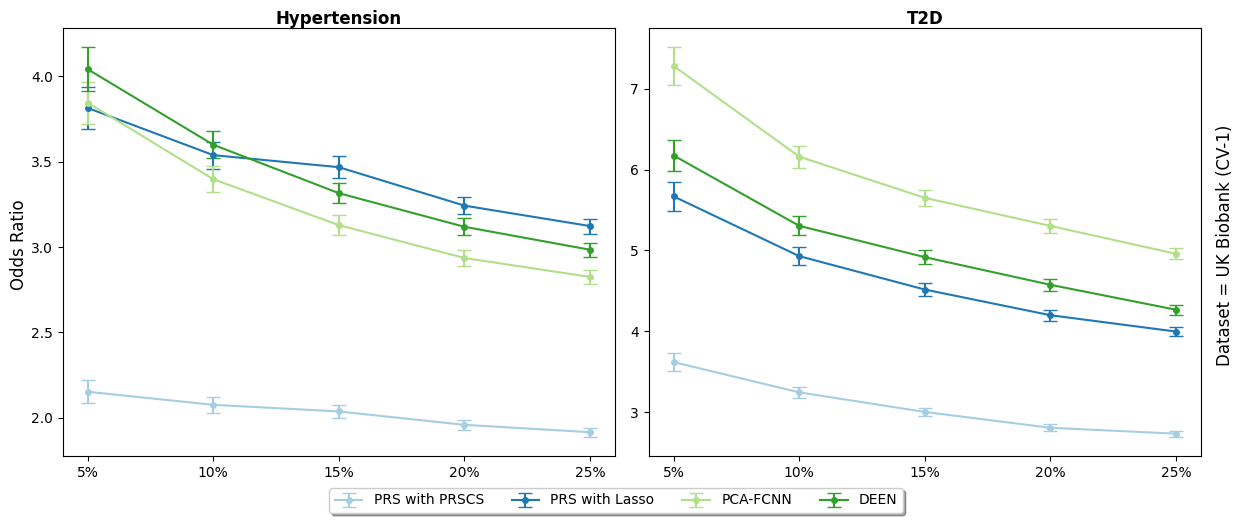

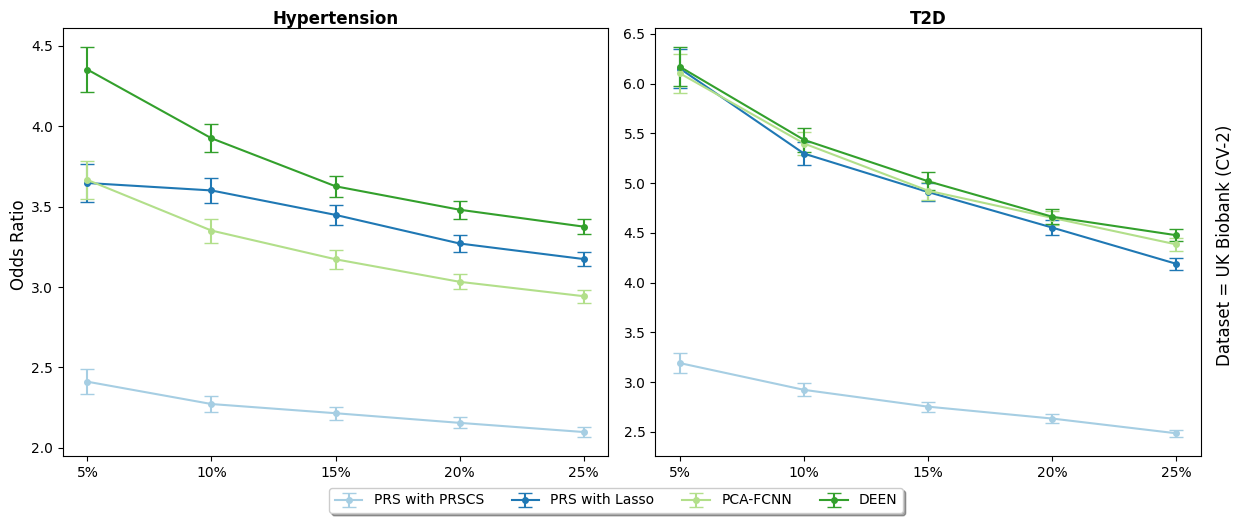

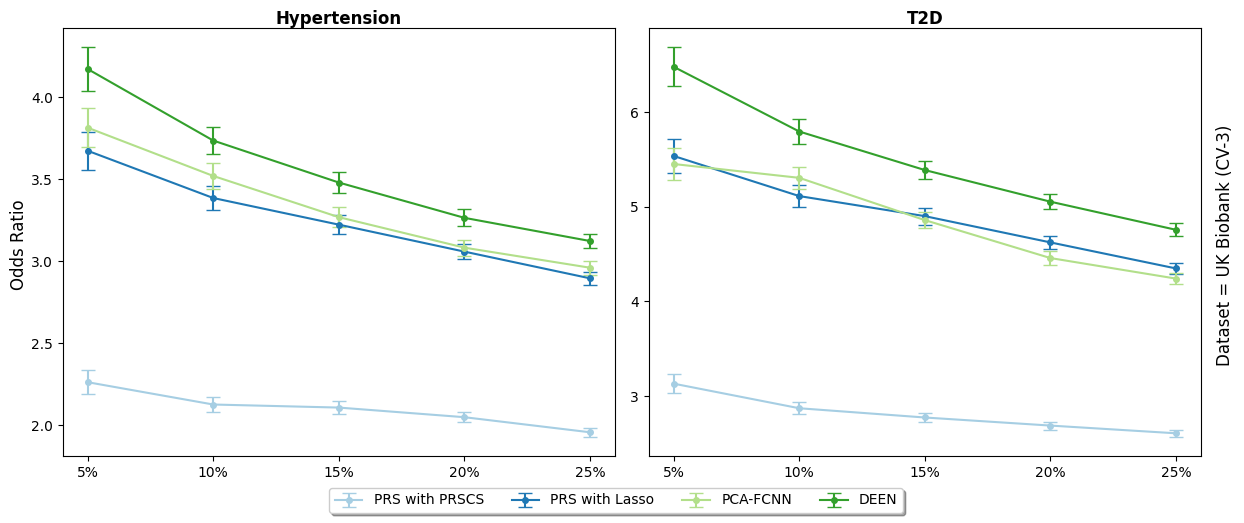

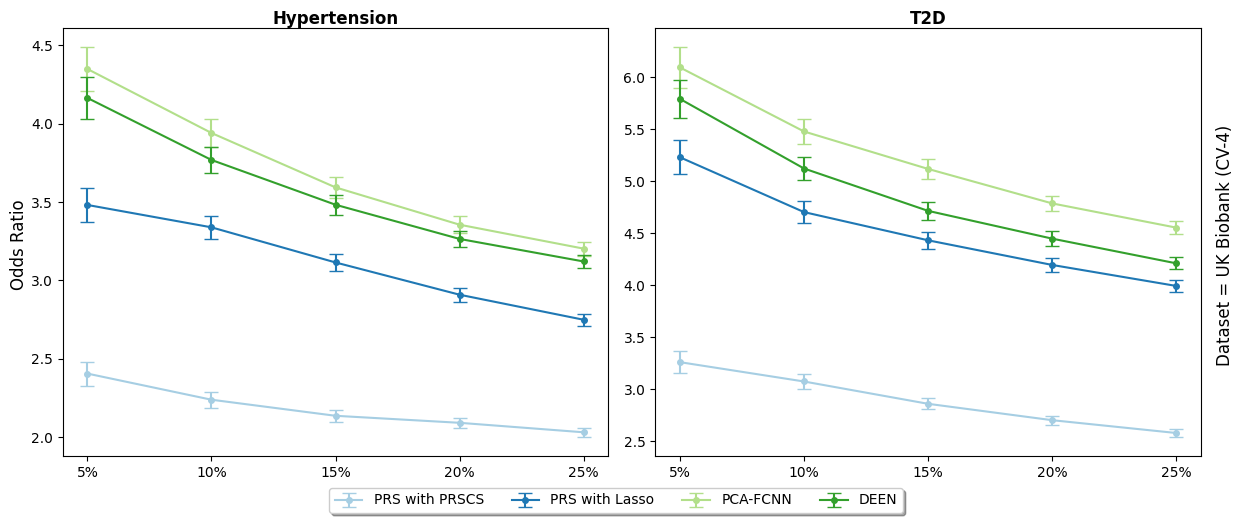

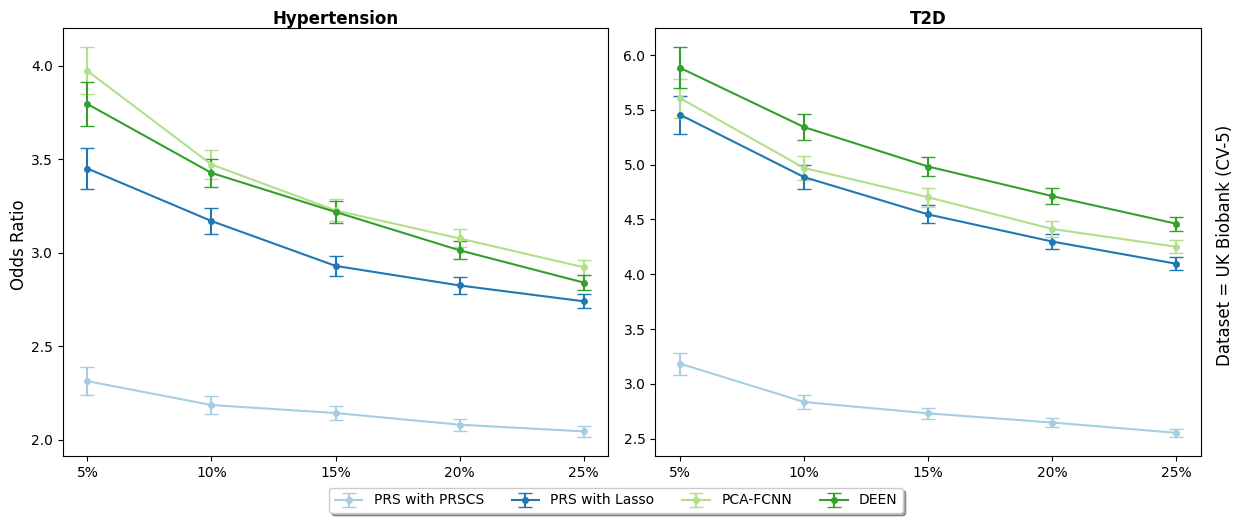

Figure S- 3 DEEN improved risk stratification for T2D and Hypertension Phenotypes for UKBB with p<0.0005 for CV-1 to CV-5. The models compared are DEEN (green), PCA-FCNN (light green), Lasso (dark blue), and PRSCS (light blue). The x-axis represents different percentage thresholds (5%, 10%, 15%, 20%, 25%) for selecting the high-risk group, and the y-axis shows the odds ratios enrichment of cases between the high- and low-risk groups for each threshold. 95% confidence intervals are shown for each odds ratio. Results for UKBB are displayed in the top row, and for AoU are on the bottom row.

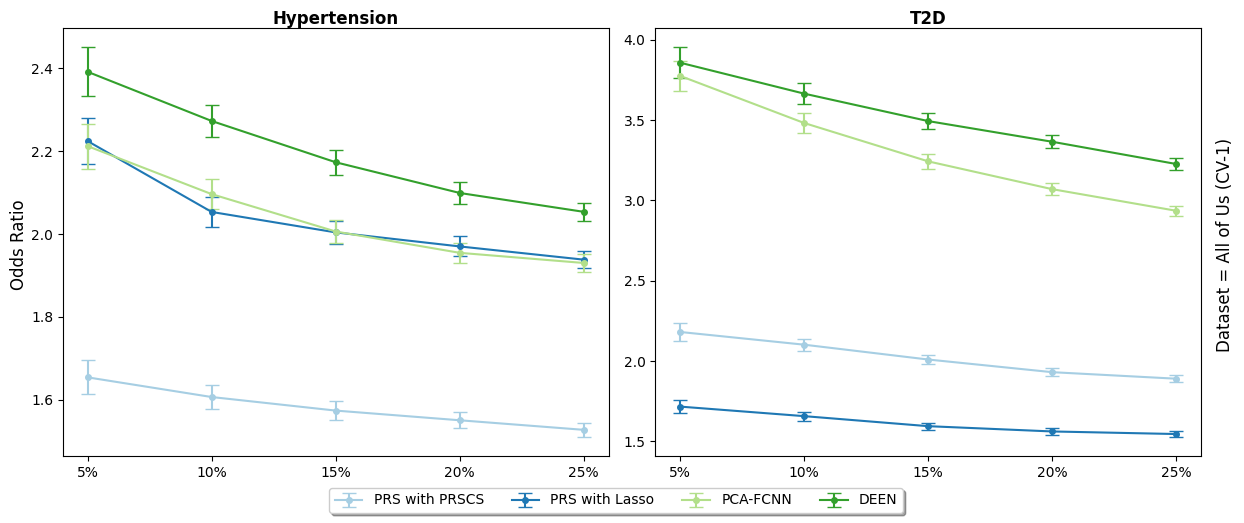

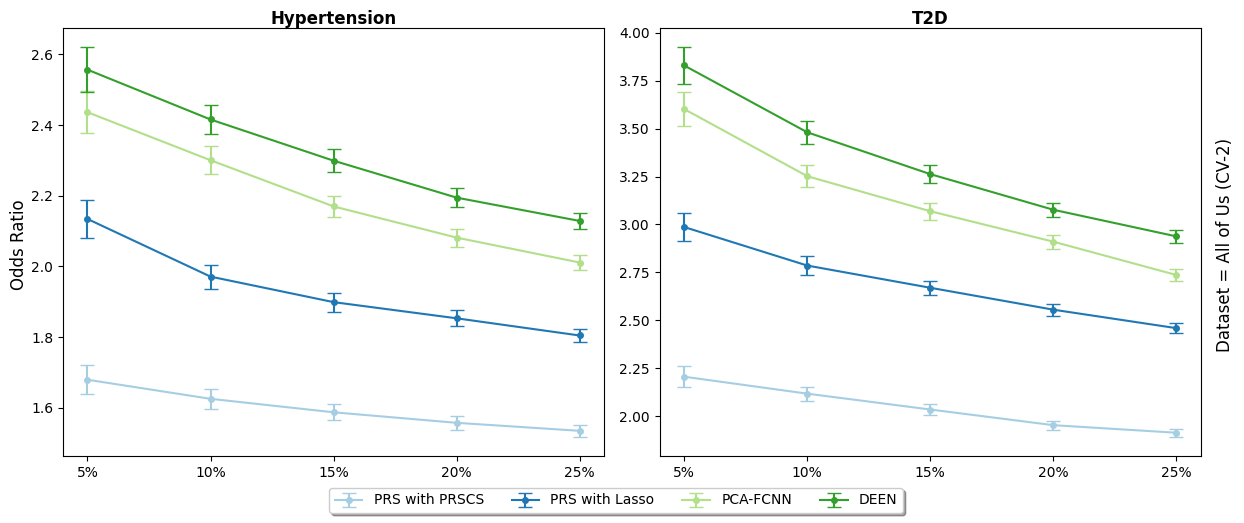

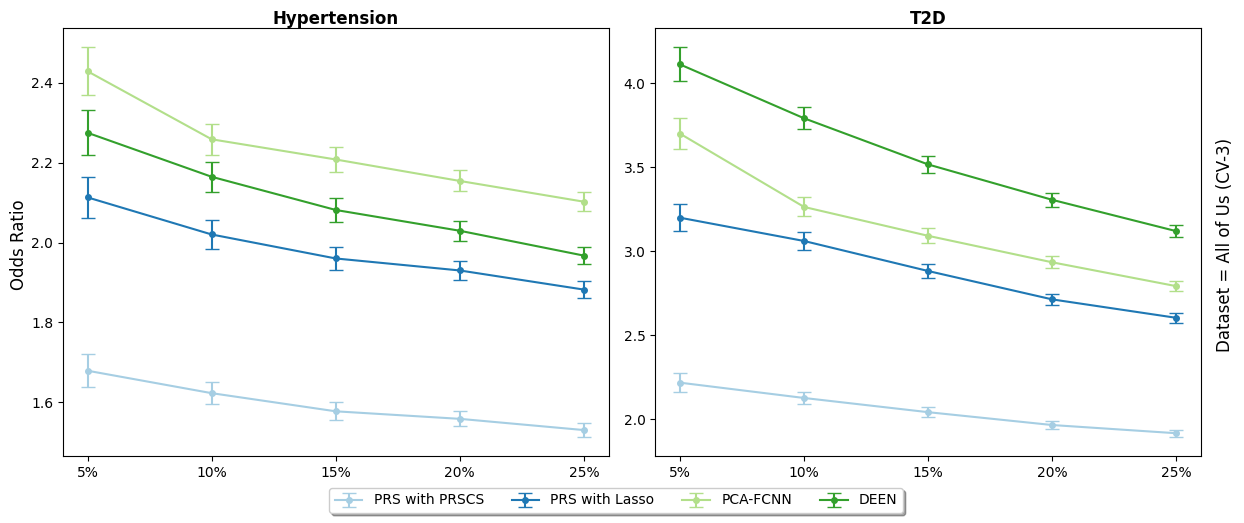

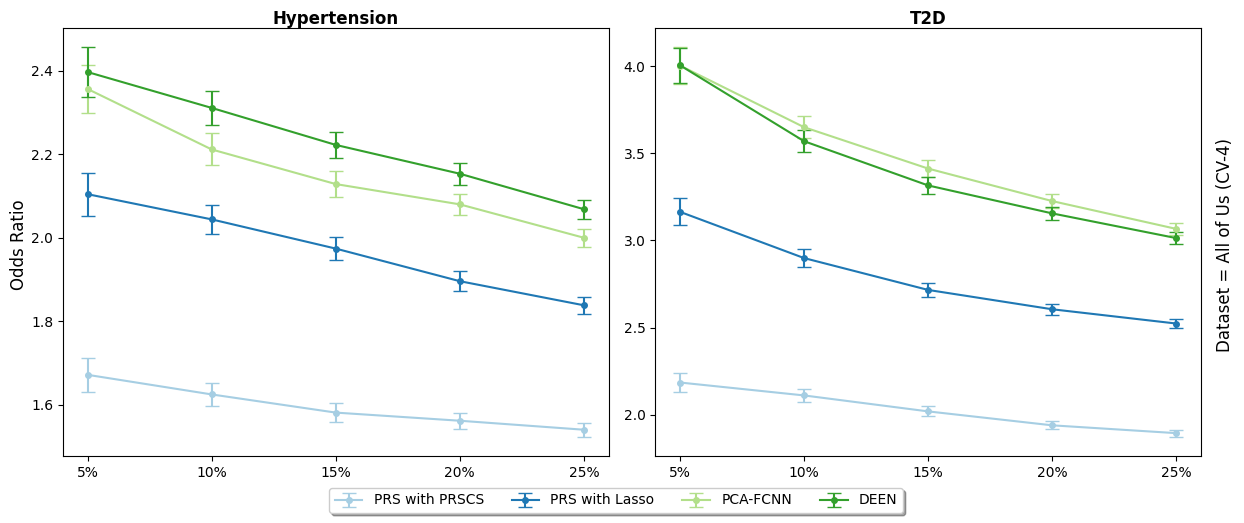

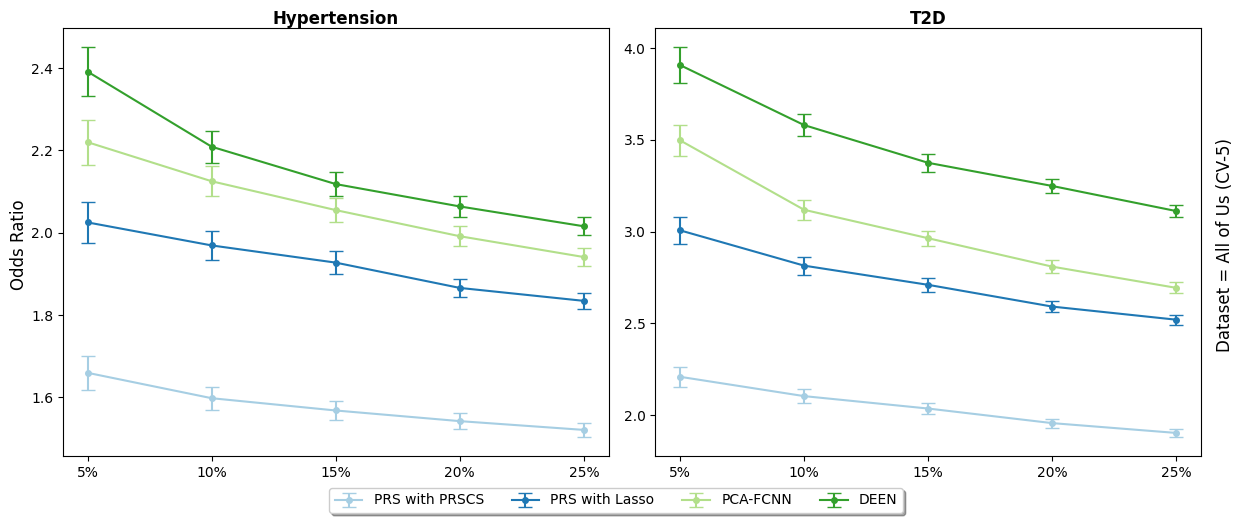

Figure S- 4 DEEN improved risk for T2D and Hypertension Phenotypes for All of Us with p<0.005 for CV-1 to CV-5. The models compared are DEEN (green), PCA-FCNN (light green), Lasso (dark blue), and PRSCS (light blue). The x-axis represents different percentage thresholds (5%, 10%, 15%, 20%, 25%) for selecting the high-risk group, and the y-axis shows the odds ratios enrichment of cases between the high- and low-risk groups for each threshold. 95% confidence intervals are shown for each odds ratio. Results for UKBB are displayed in the top row, and for AoU are on the bottom row.

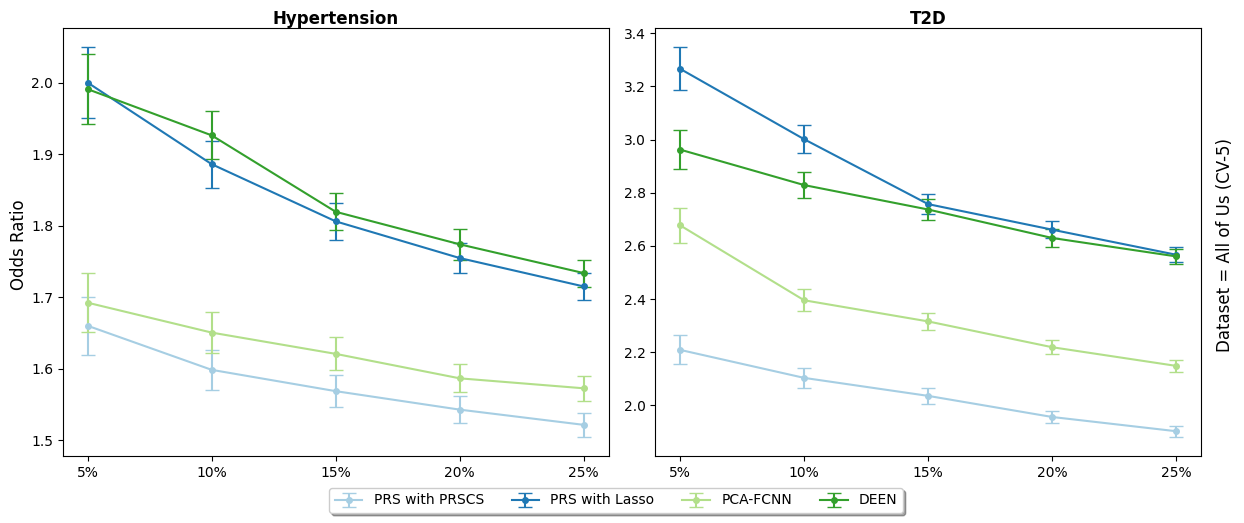

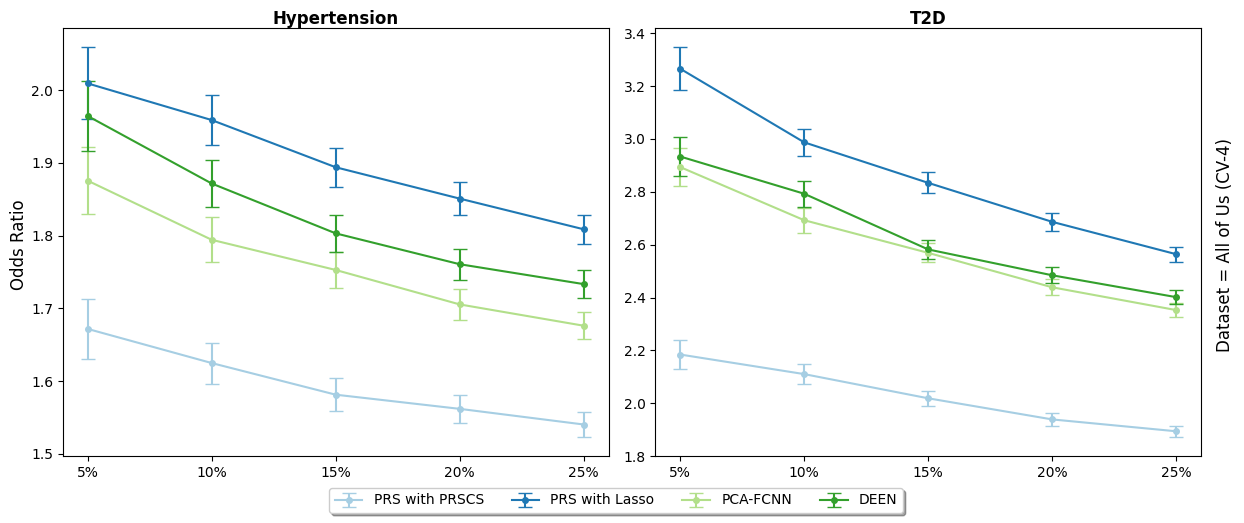

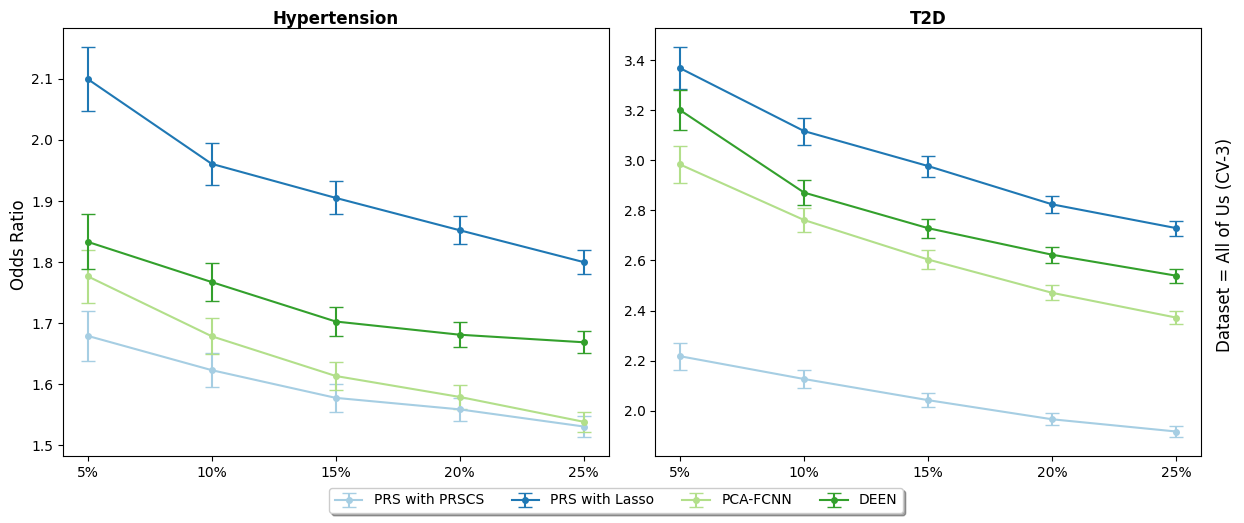

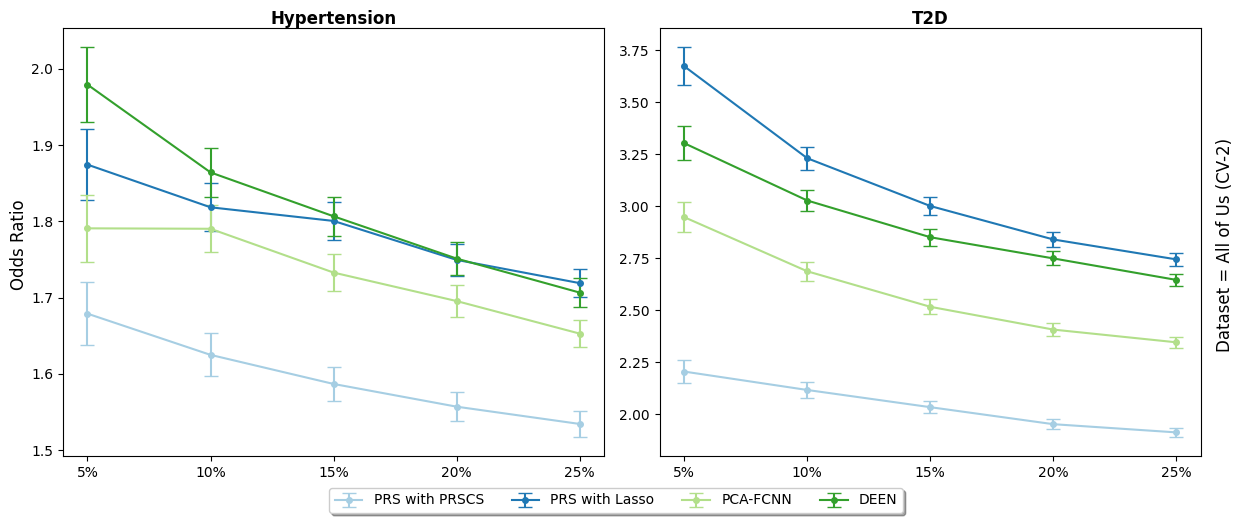

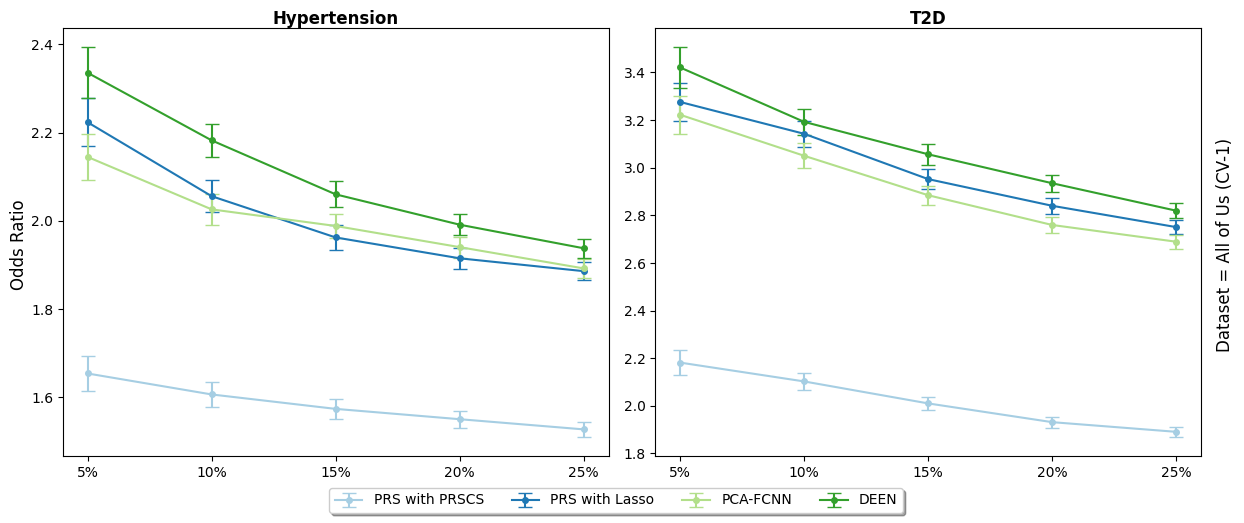

Figure S- 5 DEEN improved risk for T2D and Hypertension Phenotypes for All of Us with p<0.0005 for CV-1 to CV-5. The models compared are DEEN (green), PCA-FCNN (light green), Lasso (dark blue), and PRSCS (light blue). The x-axis represents different percentage thresholds (5%, 10%, 15%, 20%, 25%) for selecting the high-risk group, and the y-axis shows the odds ratios enrichment of cases between the high- and low-risk groups for each threshold. 95% confidence intervals are shown for each odds ratio. Results for UKBB are displayed in the top row, and for AoU are on the bottom row.

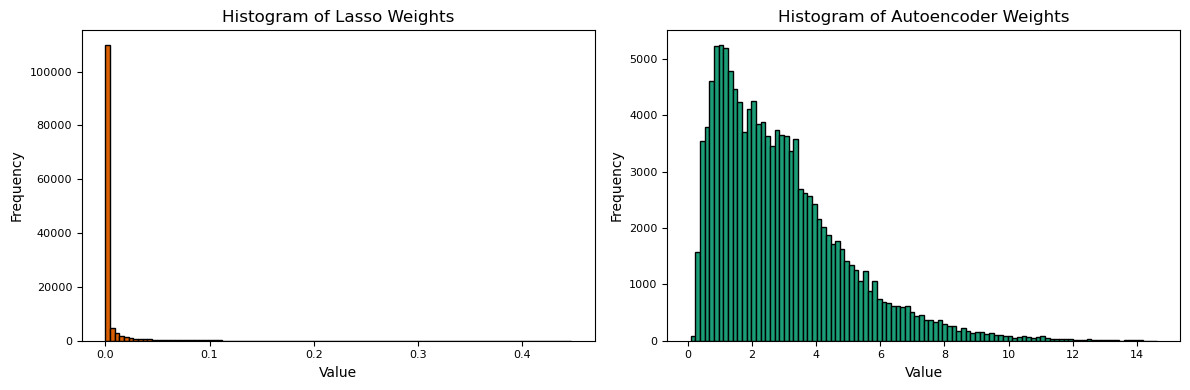

Figure S- 6 Comparison of SNP contributions between autoencoder and Lasso. Each histogram displays the frequency distribution of respective SNP weight values of Lasso regression (left) and the autoencoder model (right) for Hypertension disease.

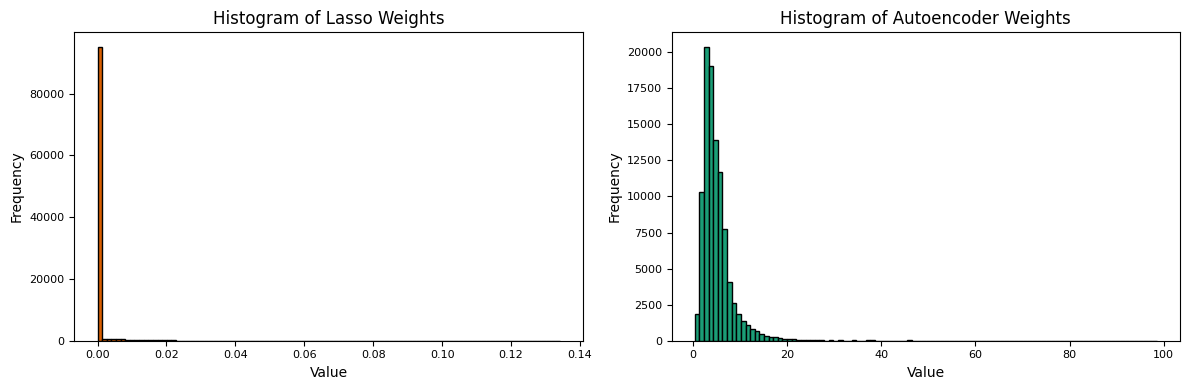

Figure S- 7 Comparison of SNP contributions between autoencoder and Lasso. Each histogram displays the frequency distribution of respective SNP weight values of Lasso regression (left) and the autoencoder model (right) for BMI disease.

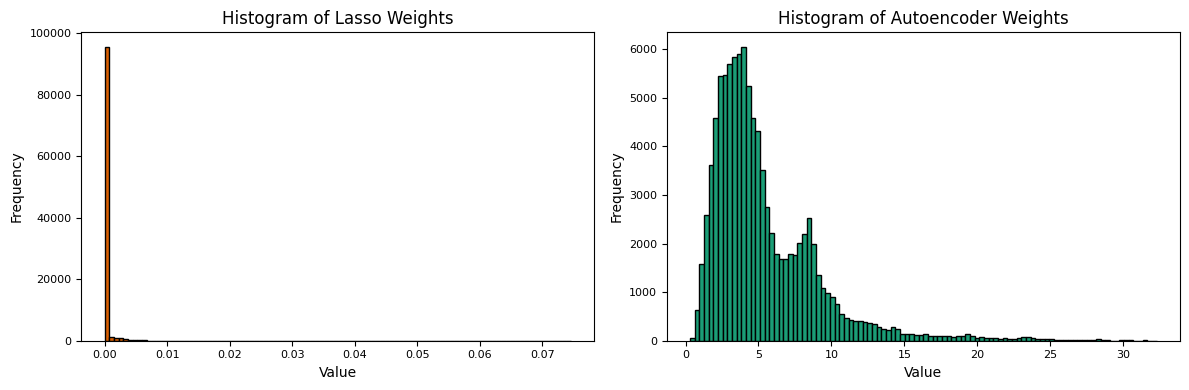

Figure S- 8 Comparison of SNP contributions between autoencoder and Lasso. Each histogram displays the frequency distribution of respective SNP weight values of Lasso regression (left) and the autoencoder model (right) for Cholesterol disease.

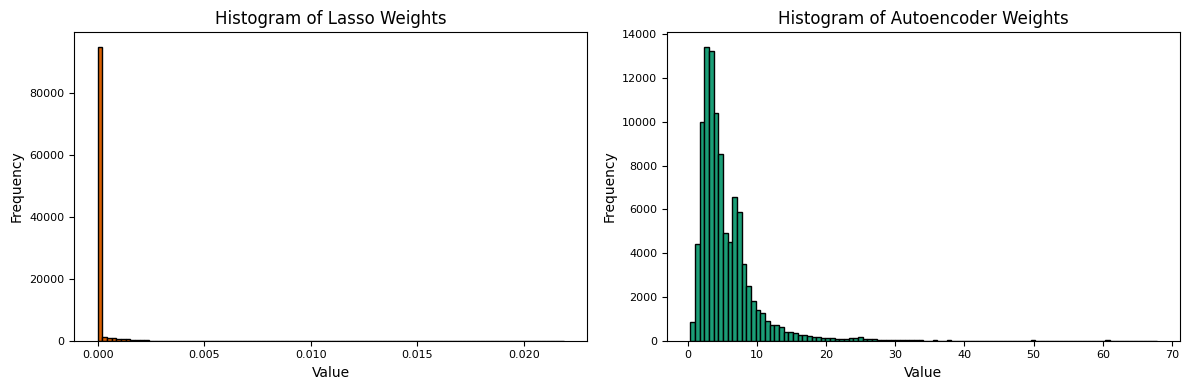

Figure S- 9 Comparison of SNP contributions between autoencoder and Lasso. Each histogram displays the frequency distribution of respective SNP weight values of Lasso regression (left) and the autoencoder model (right) for HDL disease.

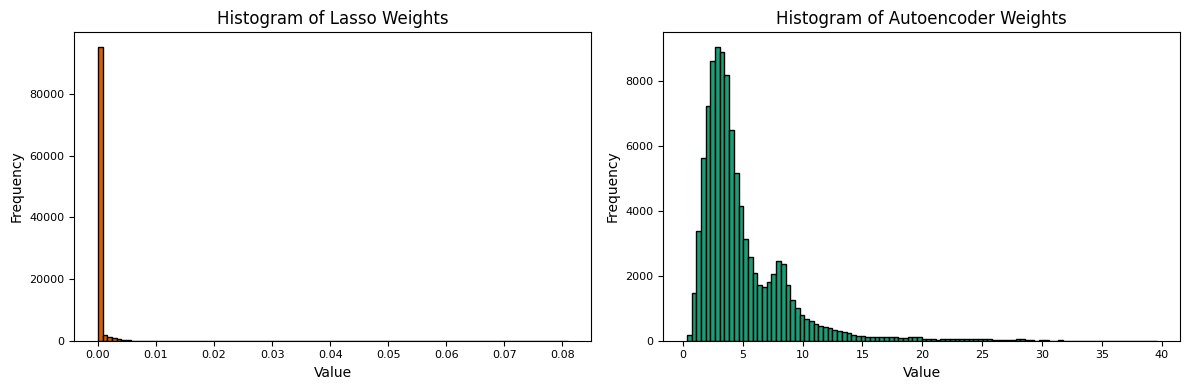

Figure S- 10 Comparison of SNP contributions between autoencoder and Lasso. Each histogram displays the frequency distribution of respective SNP weight values of Lasso regression (left) and the autoencoder model (right) for LDL disease.

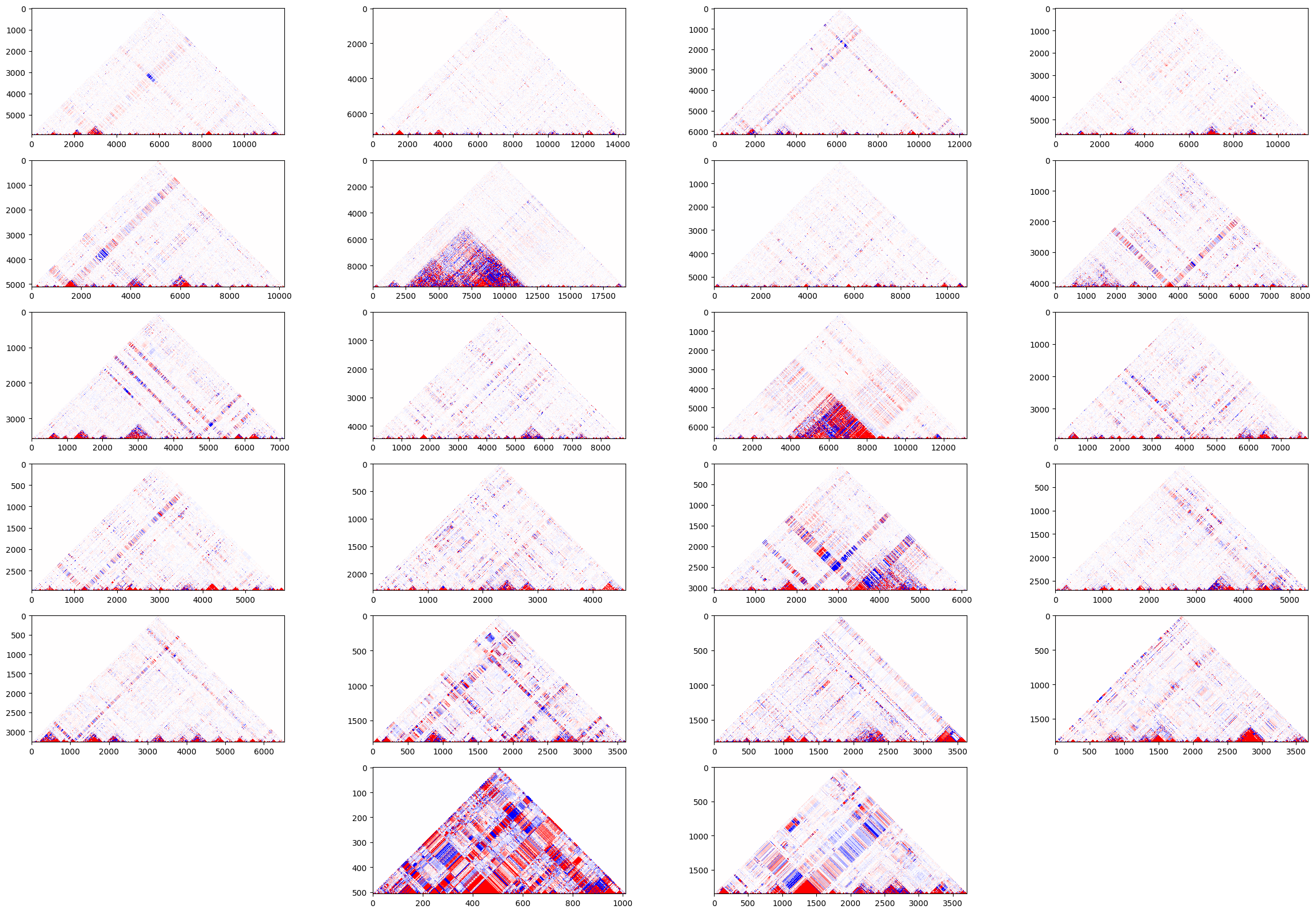

Figure S- 11 Correlations in the encoder layer nodes weights of the encoder SNP inputs, calculated using the Pearson correlation coefficient for Hypertension chromosomes 1-22. Red indicates positive correlations, while blue indicates negative correlations.

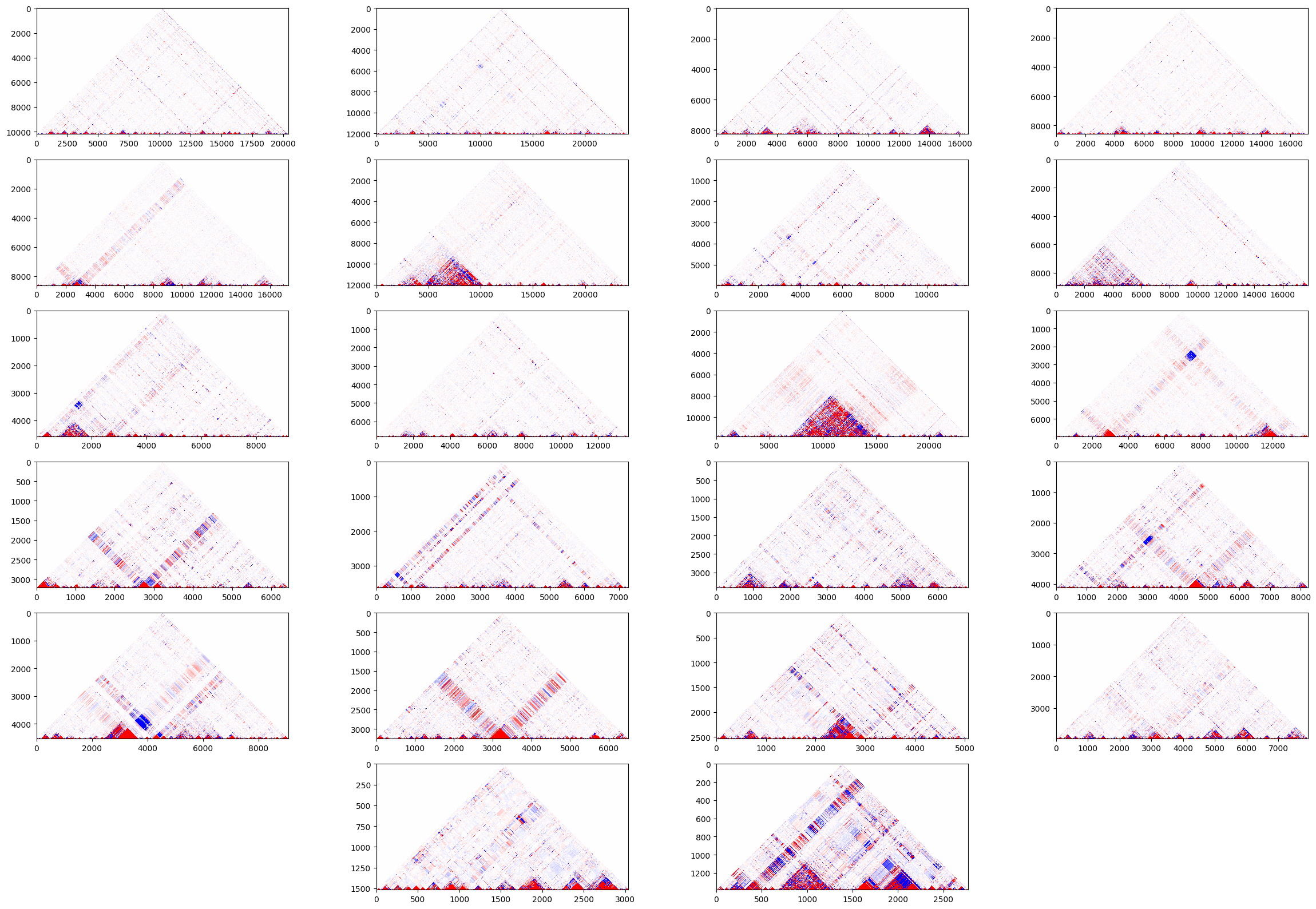

Figure S- 12 Correlations in the encoder layer nodes weights of the encoder SNP inputs, calculated using the Pearson correlation coefficient for T2D chromosomes 1-22. Red indicates positive correlations, while blue indicates negative correlations.

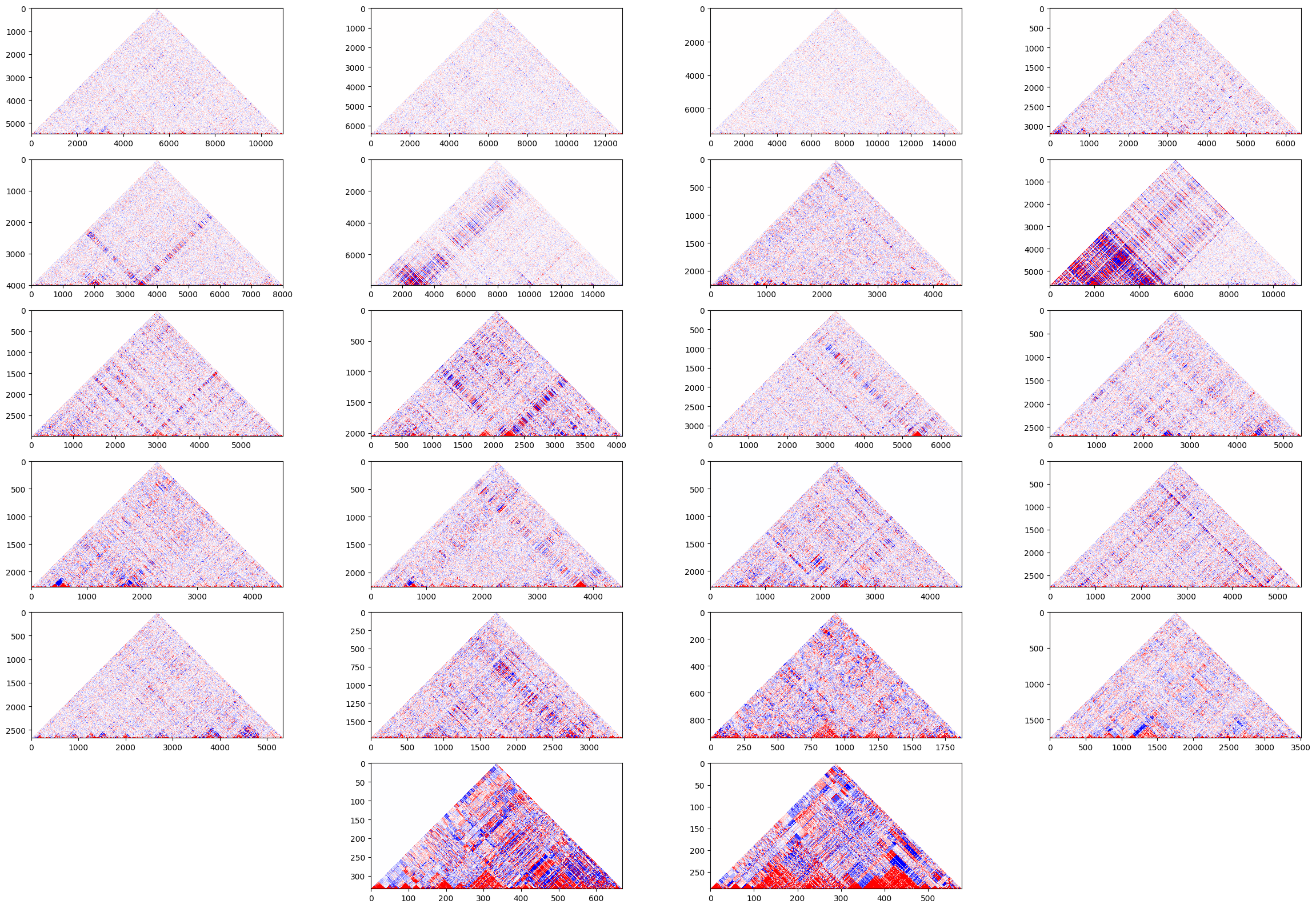

Figure S- 13 Correlations in the encoder layer nodes weights of the encoder SNP inputs, calculated using the Pearson correlation coefficient for BMI chromosomes 1-22. Red indicates positive correlations, while blue indicates negative correlations.

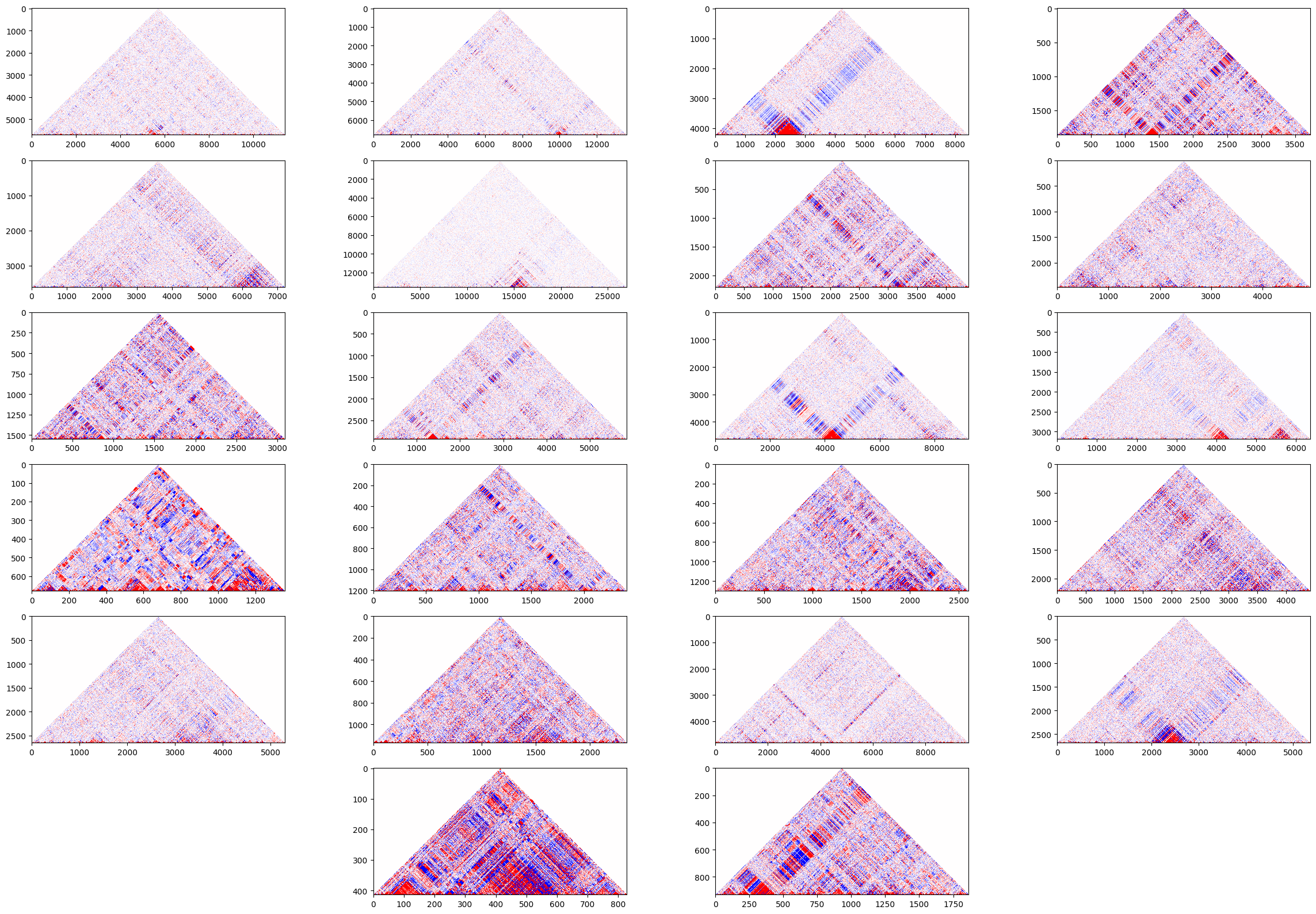

Figure S- 14 Correlations in the encoder layer nodes weights of the encoder SNP inputs, calculated using the Pearson correlation coefficient for Cholesterol chromosomes 1-22. Red indicates positive correlations, while blue indicates negative correlations.

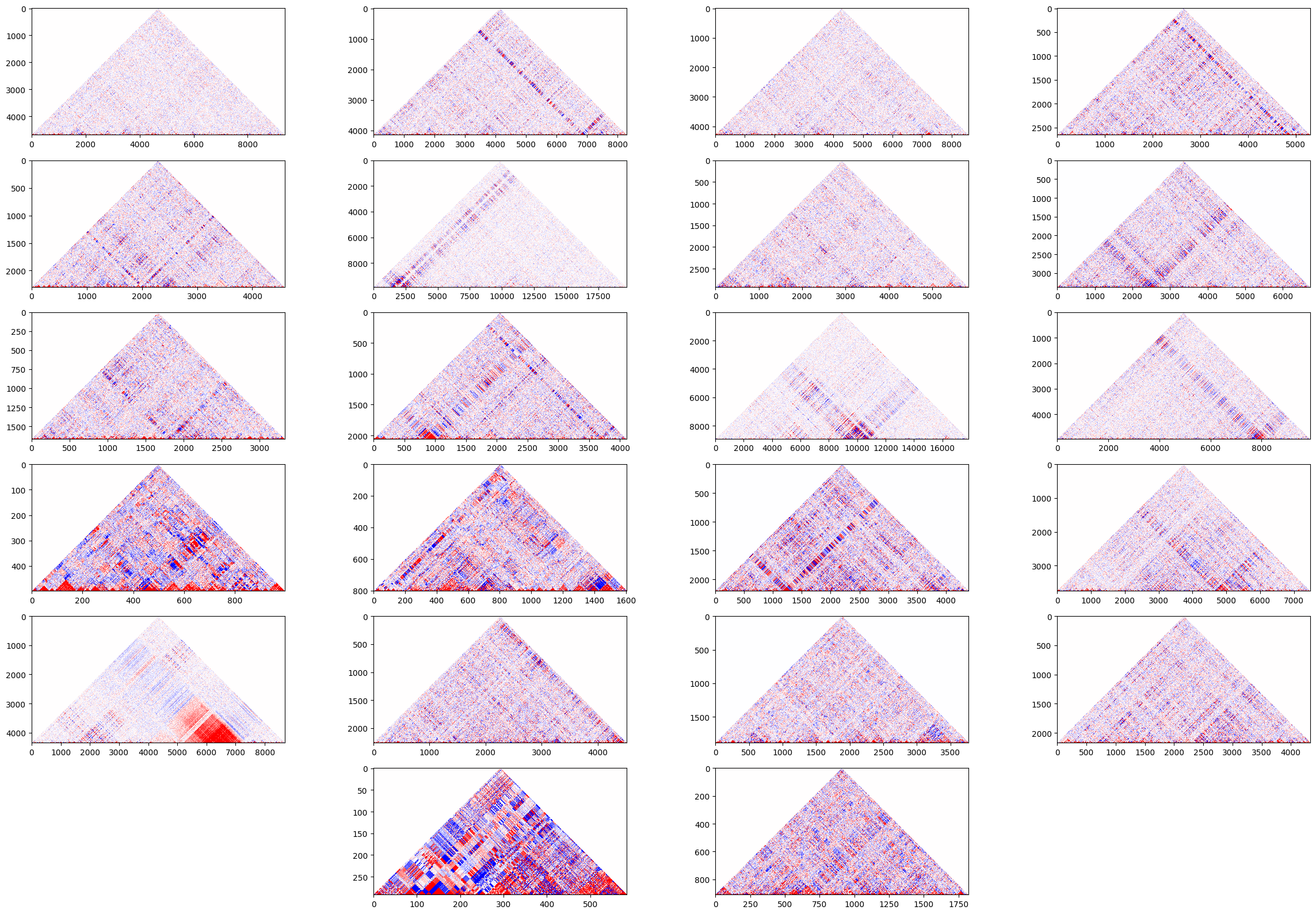

Figure S- 15 Correlations in the encoder layer nodes weights of the encoder SNP inputs, calculated using the Pearson correlation coefficient for HDL chromosomes 1-22. Red indicates positive correlations, while blue indicates negative correlations.

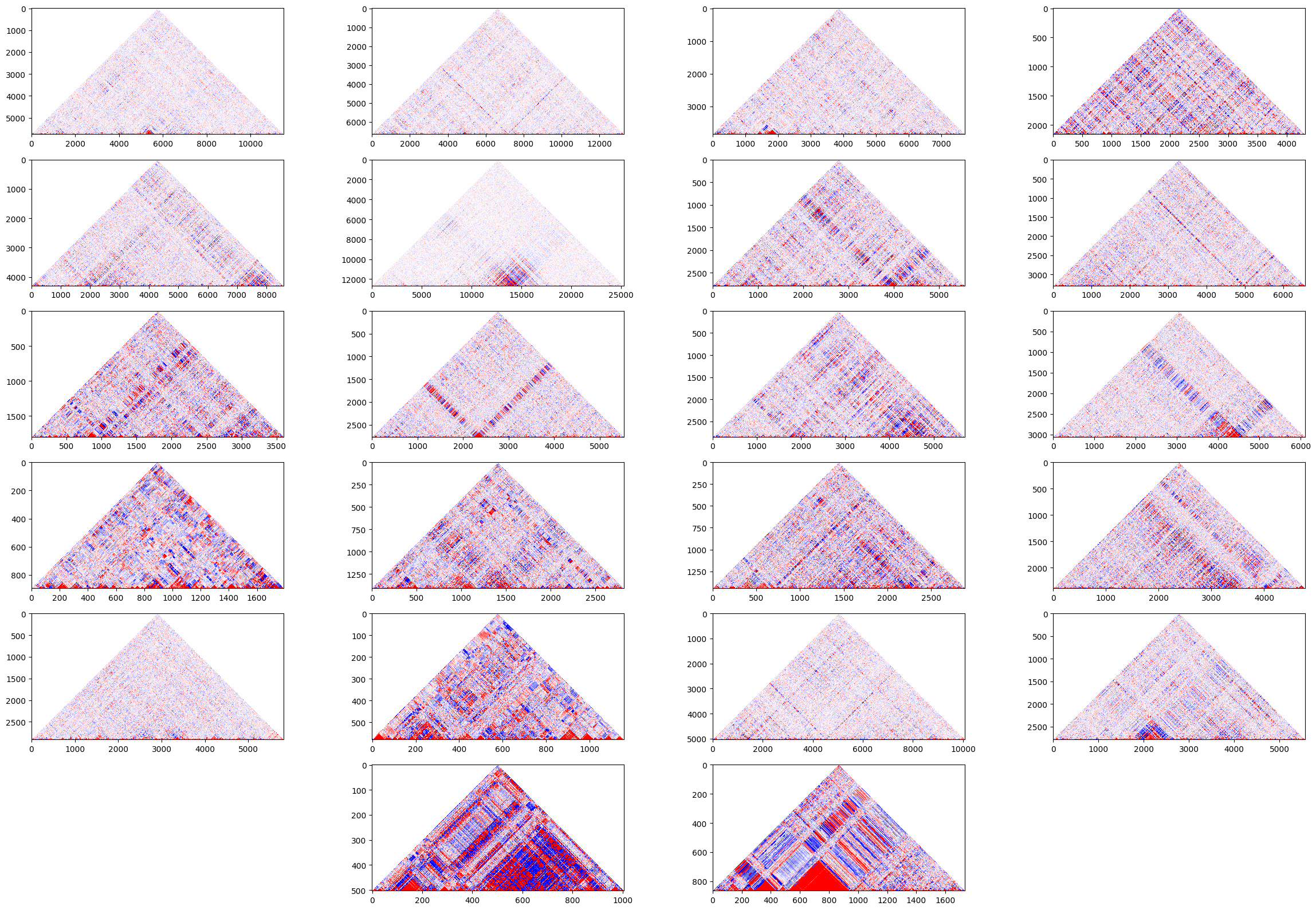

Figure S- 16 Correlations in the encoder layer nodes weights of the encoder SNP inputs, calculated using the Pearson correlation coefficient for T2D chromosomes 1-22. Red indicates positive correlations, while blue indicates negative correlations.

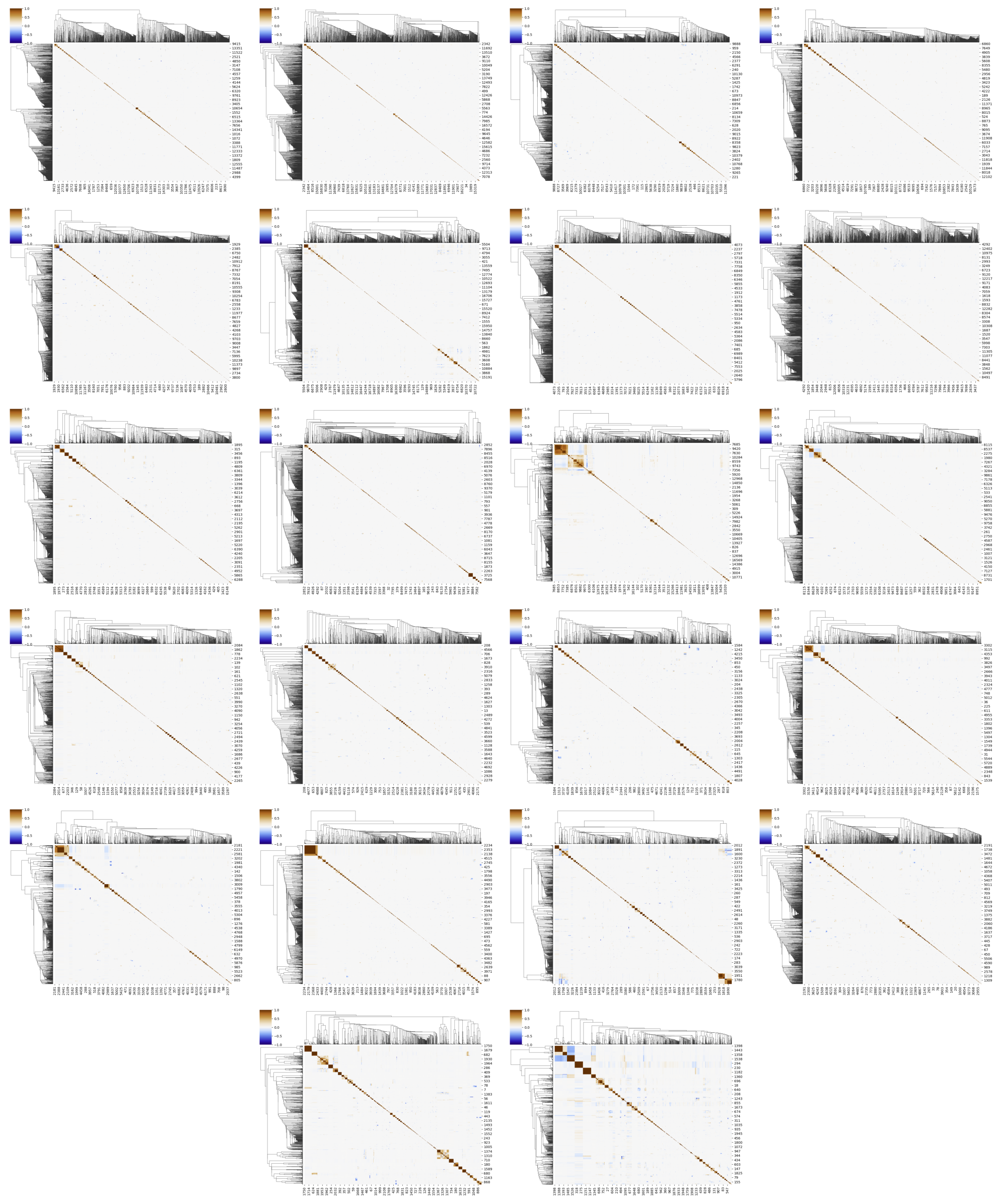

Figure S- 17 Cluster map showing the correlation values between nodes in the decoder layer of a chromosome-specific autoencoder for Hypertension, covering chromosomes 1-22. The heatmap visualizes correlations ranging from -1 to 1, with positive correlations in red, negative correlations in blue, and no correlation in white. This representation helps to identify the relationships between the nodes in the bottleneck layer weights of the encoder SNP inputs.

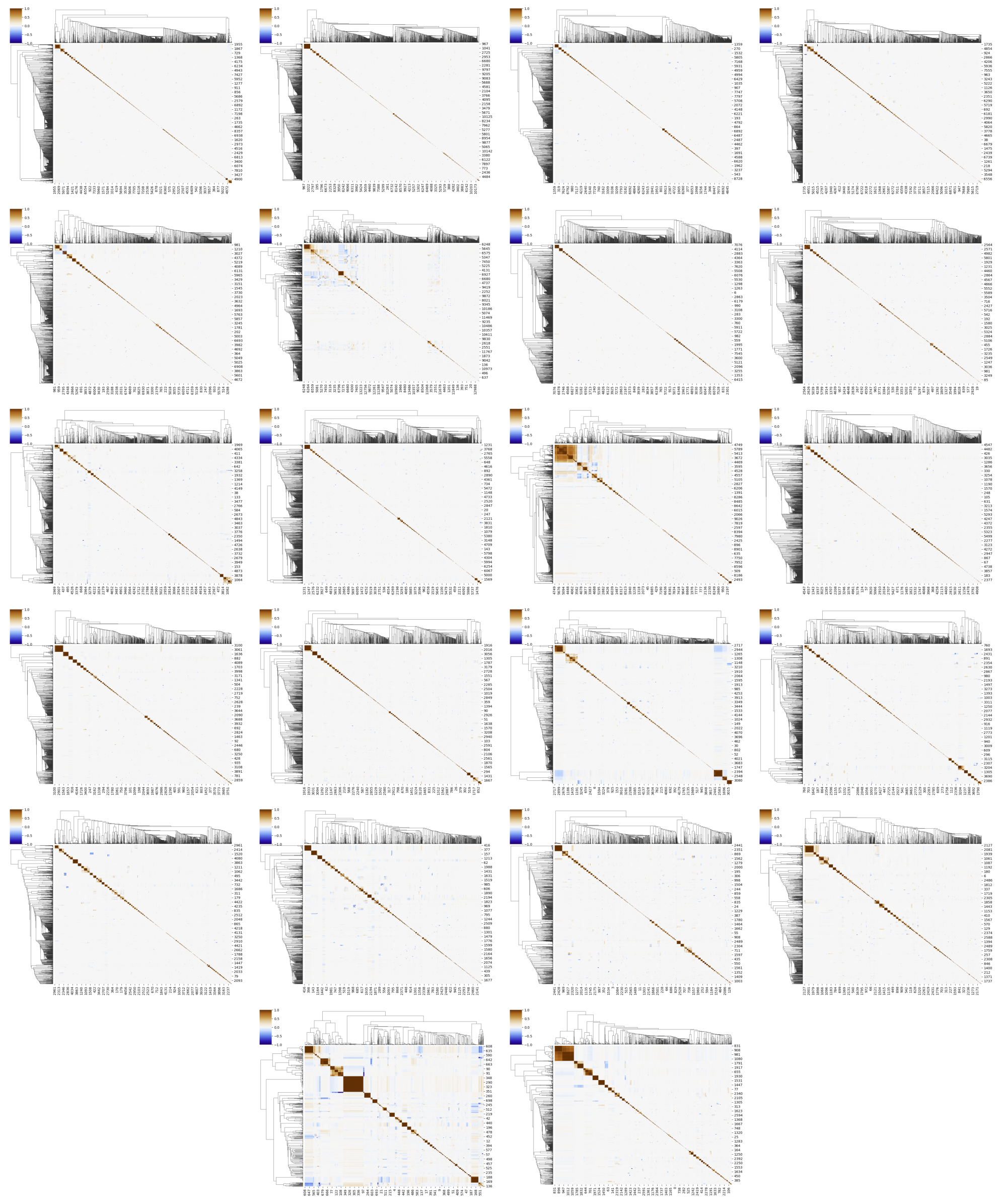

Figure S- 18 Cluster map showing the correlation values between nodes in the decoder layer of a chromosome-specific autoencoder for T2D, covering chromosomes 1-22. The heatmap visualizes correlations ranging from -1 to 1, with positive correlations in red, negative correlations in blue, and no correlation in white. This representation helps to identify the relationships between the nodes in the bottleneck layer weights of the encoder SNP inputs.

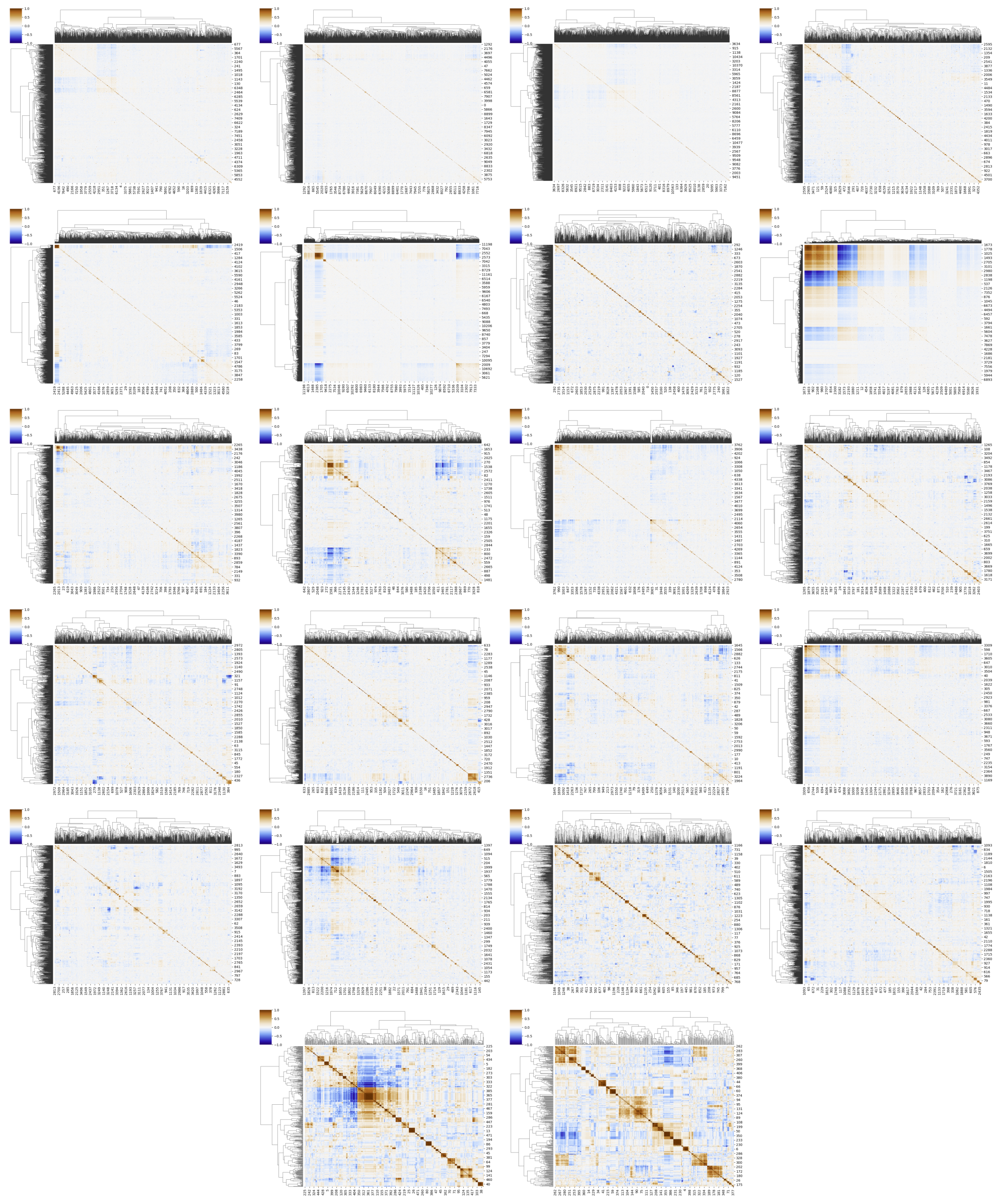

Figure S- 19 Cluster map showing the correlation values between nodes in the decoder layer of a chromosome-specific autoencoder for BMI, covering chromosomes 1-22. The heatmap visualizes correlations ranging from -1 to 1, with positive correlations in red, negative correlations in blue, and no correlation in white. This representation helps to identify the relationships between the nodes in the bottleneck layer weights of the encoder SNP inputs.

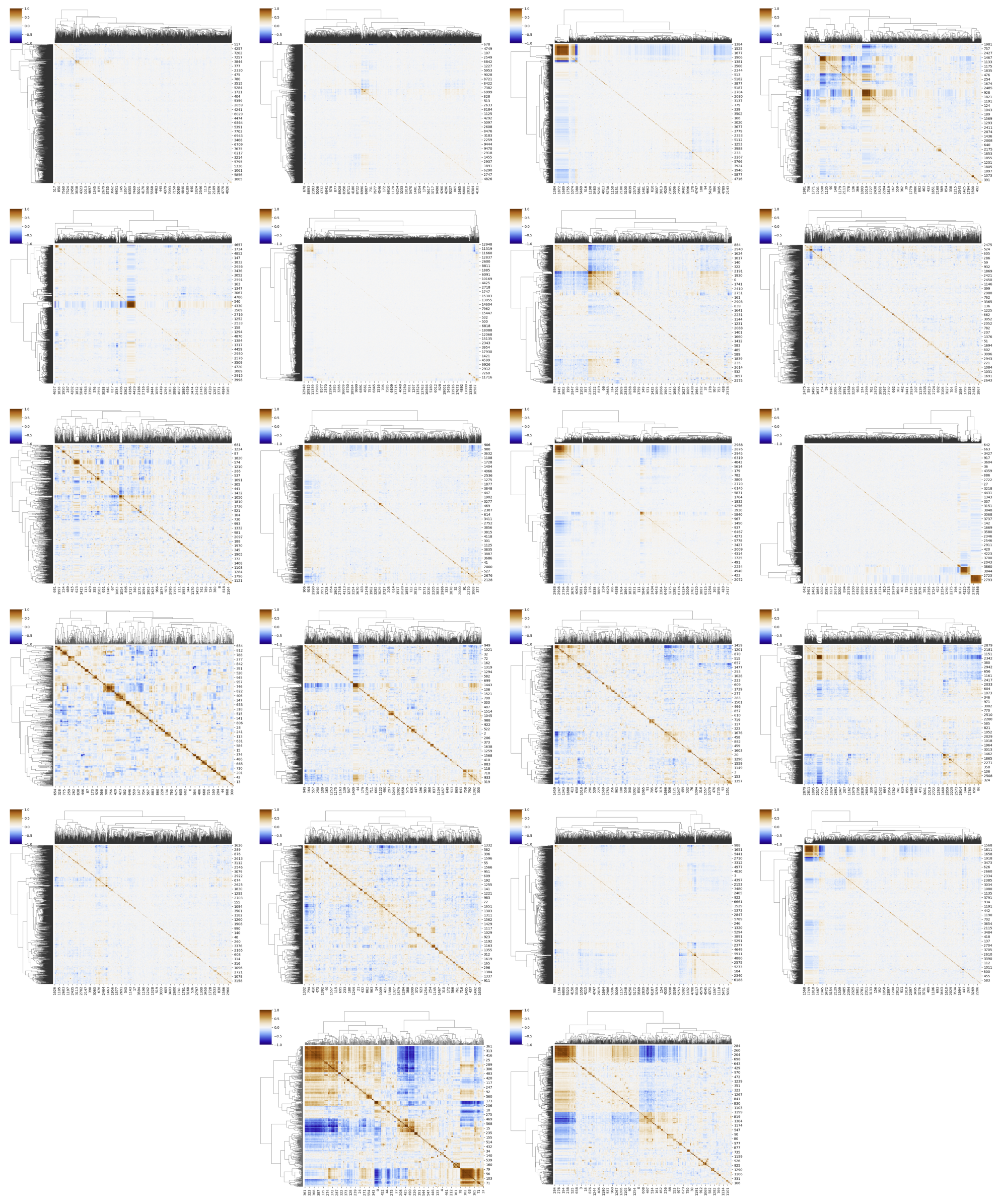

Figure S- 20 Cluster map showing the correlation values between nodes in the decoder layer of a chromosome-specific autoencoder for Cholesterol, covering chromosomes 1-22. The heatmap visualizes correlations ranging from -1 to 1, with positive correlations in red, negative correlations in blue, and no correlation in white. This representation helps to identify the relationships between the nodes in the bottleneck layer weights of the encoder SNP inputs.

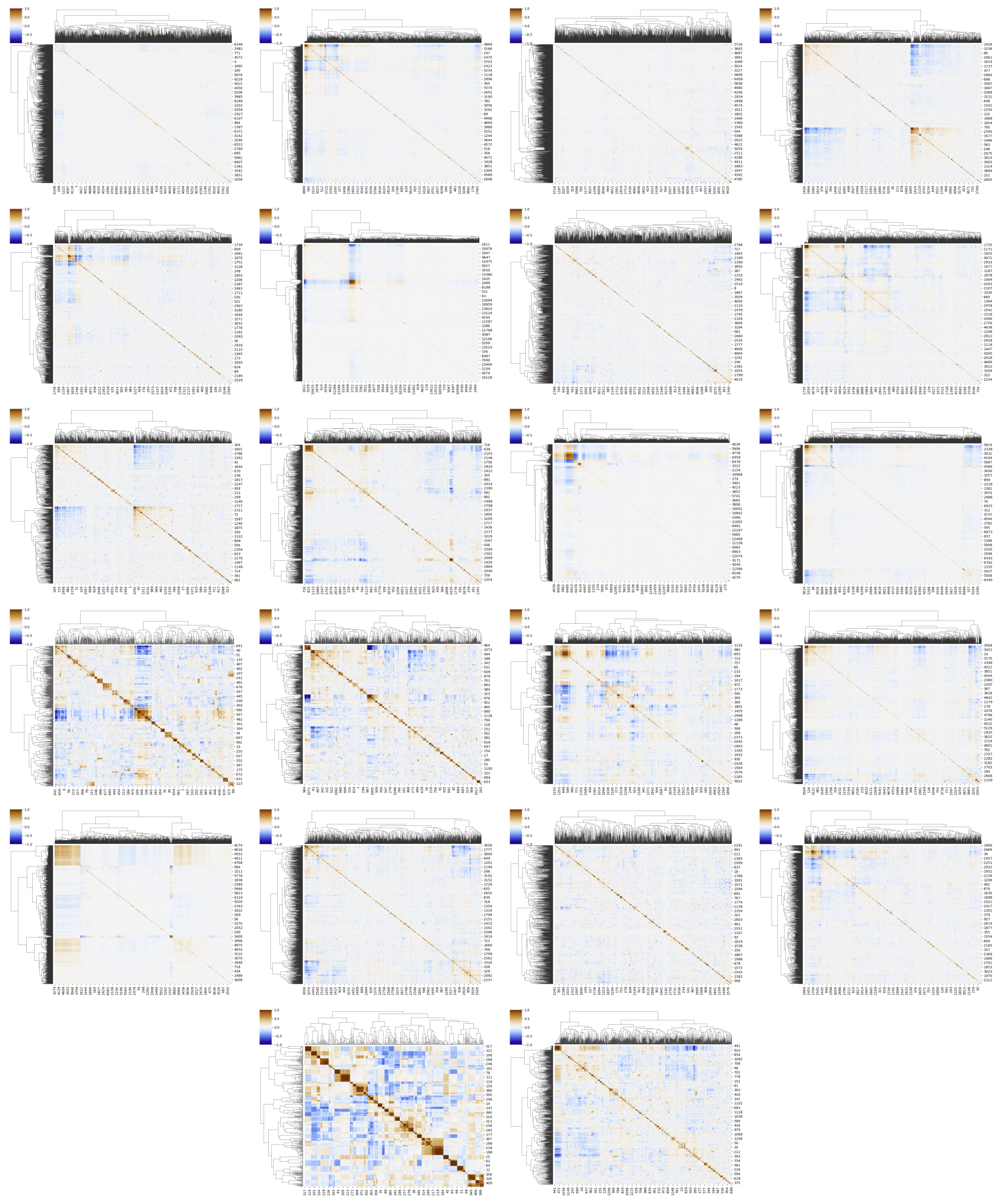

Figure S- 21 Cluster map showing the correlation values between nodes in the decoder layer of a chromosome-specific autoencoder for HDL, covering chromosomes 1-22. The heatmap visualizes correlations ranging from -1 to 1, with positive correlations in red, negative correlations in blue, and no correlation in white. This representation helps to identify the relationships between the nodes in the bottleneck layer weights of the encoder SNP inputs.

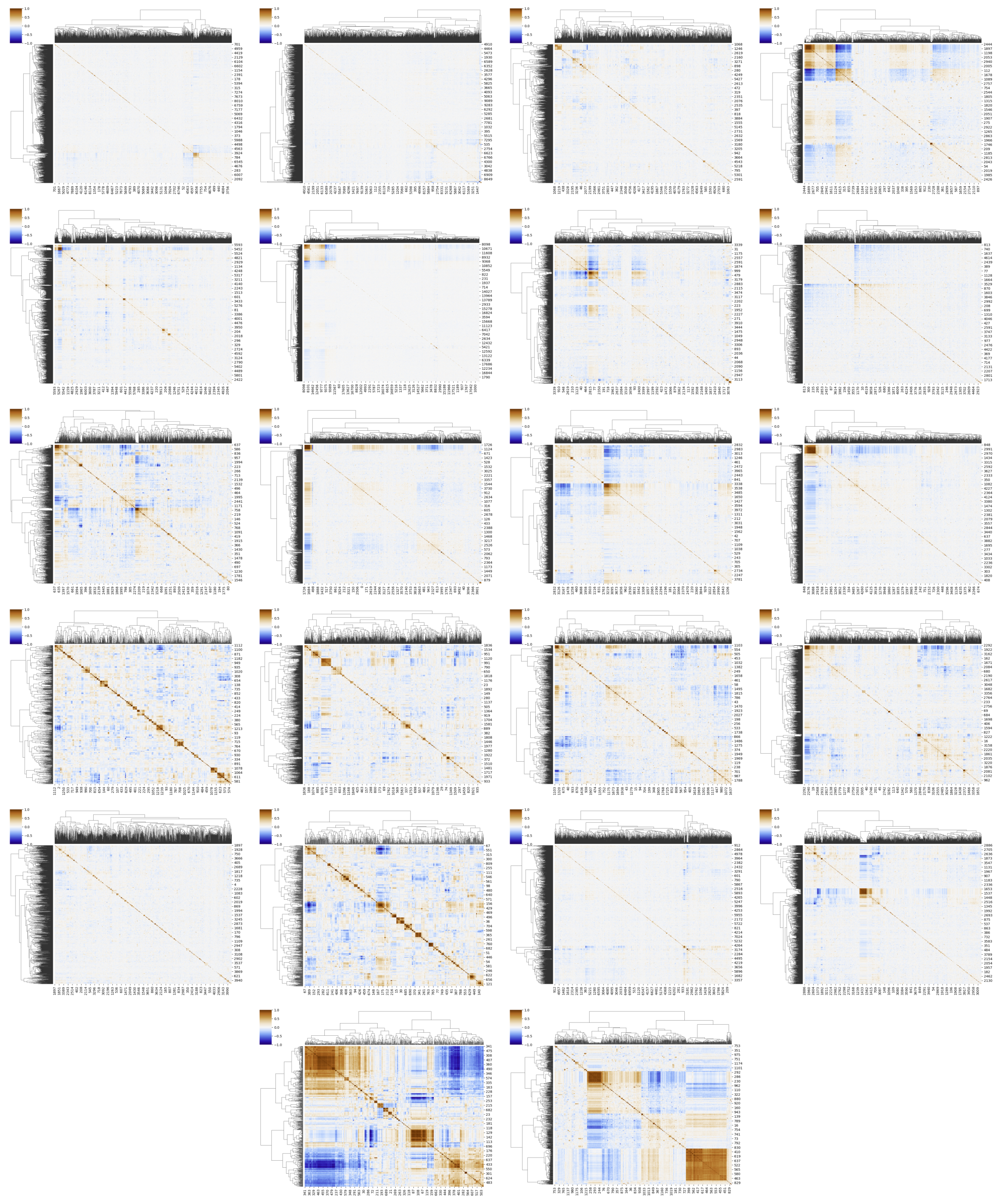

Figure S- 22 Cluster map showing the correlation values between nodes in the decoder layer of a chromosome-specific autoencoder for LDL, covering chromosomes 1-22. The heatmap visualizes correlations ranging from -1 to 1, with positive correlations in red, negative correlations in blue, and no correlation in white. This representation helps to identify the relationships between the nodes in the bottleneck layer weights of the encoder SNP inputs.
